# Feasibility of Machine Learning Analysis for the Identification of Patients with Possible Primary Ciliary Dyskinesia

**DOI:** 10.1101/2025.04.18.25326065

**Authors:** Gully Burns, Carey Kauffman, Michele Manion, Ruth-Anne Pai, Carlos Milla, Michael G. O’Connor, Adam J. Shapiro, Heidi Bjornson-Pennell

## Abstract

**BACKGROUND:** Significant diagnostic delays are common in primary ciliary dyskinesia (PCD), a rare disease that is significantly underdiagnosed. Scalable screening methods could improve early identification and health outcomes.

**RESEARCH QUESTION:** Can machine learning (ML) be used to screen for PCD in pediatric patients?

**STUDY DESIGN AND METHODS:** We evaluated the feasibility of a random forest model to screen for PCD using data from the PCD Foundation Registry and a national claims database. We identified a cohort of pediatric patients with diagnostic codes indicative of conditions potentially associated with PCD, and studied diagnostic, procedural, and pharmaceutical codes associated with PCD to develop ML features. Models were trained on composite claims data from confirmed patients with PCD, patients with Q34.8 (Specific Congenital Malformation of the Respiratory System) diagnosed within six months of an Electron Microscopy procedure (Q34.8+EM), and a randomly-selected, matched control group. Model performance was tested through 5-fold cross-validation.

**RESULTS:** Using 82 confirmed PCD cases and 4,161 matched controls, the model demonstrated variable performance (positive predictive value 0.45–0.73, sensitivity 0.75–0.94). Synthetic data augmentation did not improve results (positive predictive value 0.45–0.67, sensitivity 0.71–1.00). Expanding the dataset to include 319 Q34.8+EM patients and 8,214 controls improved performance (positive predictive value 0.51–0.54, sensitivity 0.82–0.90), suitable for screening. In a cohort of 1.32 million pediatric patients, 7,705 were classified as positive, consistent with the estimated prevalence of PCD (1:7,554).

**INTERPRETATION:** This study demonstrates the feasibility of using ML to screen for PCD using claims data, even in the absence of a specific International Classification of Disease (ICD) code. Such screening approaches may aid in the identification of individuals who may benefit from timely diagnostic testing and targeted interventions.

## INTRODUCTION

Primary ciliary dyskinesia (PCD) is a rare, progressive genetic disorder affecting the function of motile cilia. Dyskinetic ciliary movement results in stagnant mucus throughout the upper and lower airways, causing recurrent ear, sinonasal, and pulmonary infections that eventually result in bronchiectasis and may lead to respiratory failure requiring lung transplantation. Patients may also present with left-right organ laterality defects, subfertility, and neonatal respiratory distress^1,2^. The prevalence of PCD is estimated to be at least 1 in 7,554, but is likely underdiagnosed due to barriers such as lack of awareness and phenotypic overlap with common respiratory diseases^3,4^.

Although an estimated 45,000 individuals in North America are living with PCD, about 1,000 have been diagnosed. Clinical symptoms of PCD often appear early in life, yet many cases remain undiagnosed until later childhood or even through adulthood. International communities face additional significant disparities in access to diagnosis and care^25^. This diagnostic delay significantly affects patients and their families. A timely diagnosis offers access to community support from the PCD Foundation (PCDF), enables targeted therapies, and helps avoid unnecessary or harmful treatments. At a broader level, increasing diagnosis rates will facilitate more comprehensive studies of PCD phenotypes and genotypes, and provide a stronger foundation for future clinical trials.

Several factors contribute to the delayed and underdiagnosis of PCD. There is no single “gold standard” test for diagnosing all forms of PCD^6^. Current guidelines from the American Thoracic Society^7^ and European Respiratory Society^8^ recommend genetic testing and/or ciliary electron microscopy, but these tests fail to confirm 20–30% of cases. Additional tests, such as nasal nitric oxide measurement, high-speed videomicroscopy of ciliary beat patterns, and ciliary protein immunofluorescence, are available at select centers but are not definitively diagnostic on their own^7-9^. These limitations lead to significant diagnostic delays, preventing timely referral for specialized testing and treatment, which may mitigate long-term respiratory damage and morbidity.

Although clinical screening criteria exist for PCD, they are rarely applied in primary care settings where earlier diagnosis is crucial. A recent study highlighted this challenge, where researchers employed keyword searches of electronic medical records at a single institution to identify missed PCD cases^10^. From a pool of 874 records, 21 individuals exhibited sufficient symptoms to warrant screening, ultimately leading to four PCD diagnoses. This underscores the need for improved PCD detection through review of medical records, which is complicated by the lack of a specific International Classification of Disease (ICD) code for PCD. Automated screening tools, particularly those leveraging machine learning (ML), offer a promising solution that, once validated, could be deployed in diverse clinical settings. ML has proven successful in developing screening tools for other rare diseases, with several already implemented clinically^11-15^.

In this work, we evaluate the potential of developing a predictive model using data from the PCD Foundation Registry (PCDFR) and national insurance claims to identify patients with possible PCD. We also address the key technical challenge of applying ML to rare disease screening by using patient data at scale for broad coverage combined with the deidentified identification of a positive diagnosis group for model training.

## METHODS AND MATERIALS

### Study Design and Population

In collaboration with data scientists, PCD clinician experts, and patient advocates, we developed a screening tool for PCD using datasets from the PCDFR and the Komodo Healthcare Map claims database (**Figure 1**). Diagnostic codes associated with PCD were used to query the claims database and define a screening cohort for model development and evaluation (**e-Appendix 1**). Confirmed pediatric PCD cases from the PCDFR were linked to the claims database using privacy-preserving record linkage via Datavant^16^. A background class of presumed PCD-negative cases was created from the screening cohort, matched to the positive class by age, sex, and ZIP-3. Diagnosis, procedure, and drug codes (**e-Appendix 2**) were captured and scored for all encounters in both classes. We trained a random forest model to classify patients most likely to have PCD based on symptoms and procedures associated with the disease and evaluated the model’s performance. The setting of this study is not restricted to any one facility (or even type of facility) since the health insurance records being used were generated by medical encounters across settings.

**Figure 1.**
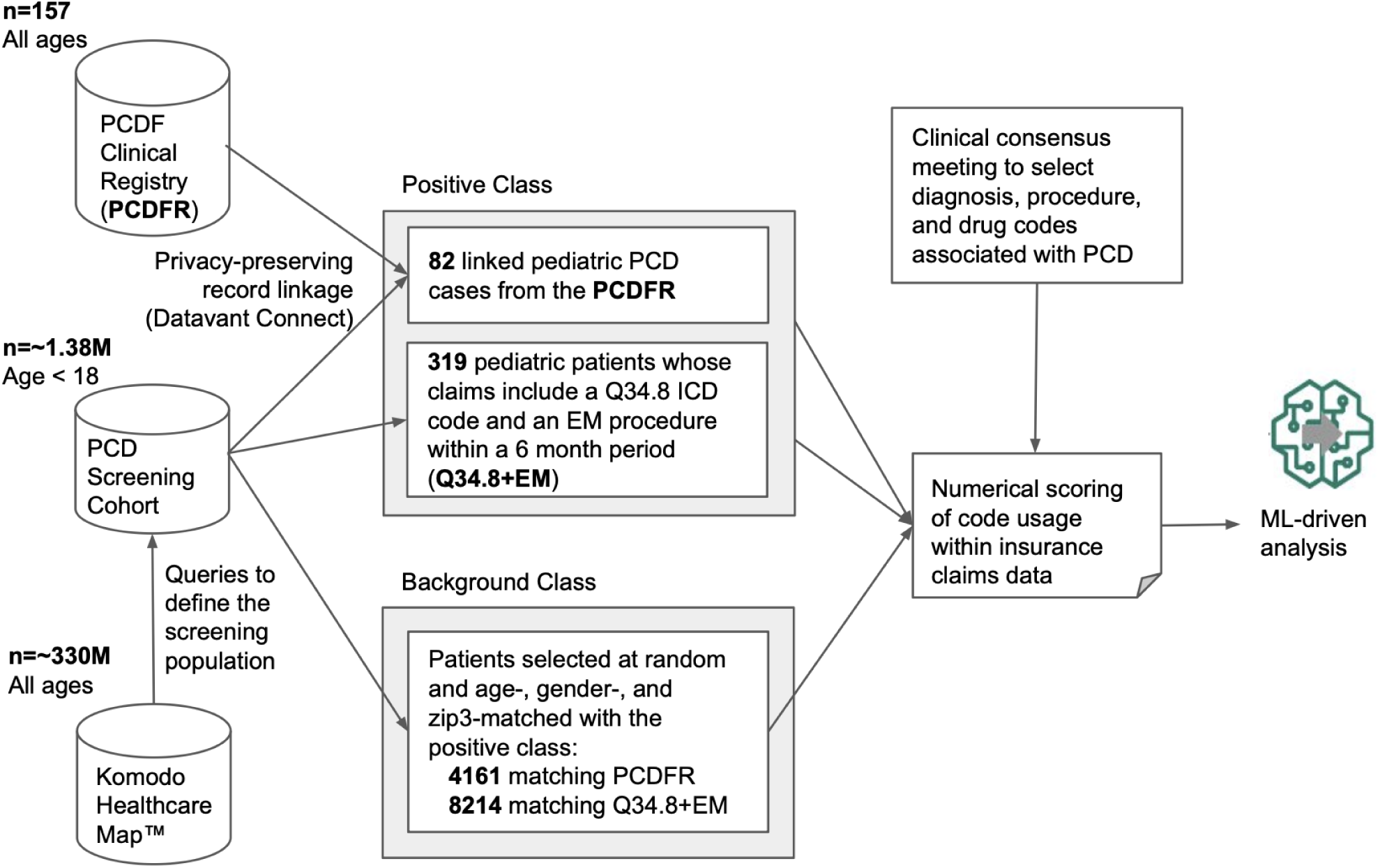
Workflow for data capture and preparation. Using a commercial privacy-preserving method, 82 confirmed pediatric patients with PCD from the PCDFR were identified within the insurance claims database and annotated as a subcohort of the positive class. Next, 319 pediatric patients whose claims include a Q34.8 ICD code and an EM procedure code within a 6 month period were identified within the insurance claims database and included as a second sub-cohort within the positive class. To generate a background cohort of pediatric patients on which to screen for primary ciliary dyskinesia, we first identified approximately 1.38M pediatric patients with insurance claims codes associated with PCD and then identified 12,375 pediatric patients matched to the positive class by age, gender, and zip3 codes. In collaboration with PCD clinical experts, we studied diagnostic, procedural, and pharmaceutical codes for patients with PCD to develop a classification model on which to perform numerical scoring of code usage within insurance claims data in the claims dataset. The classification model and numerical scores from the positive class and background class were then utilized for downstream ML analysis.

### Screening cohort

We created a screening cohort using Komodo Health’s Healthcare Map™, a nationally representative, de-identified insurance claims database. Claims codes associated with PCD diagnostic criteria and clinical features (**e-Appendix 1**) were grouped into Broad Feature Categories, which informed the queries used to generate the cohort (**Table 1**). To identify possible PCD cases, we required common symptom, diagnostic, or therapeutic codes to appear at least nine times over 18 months. No distinction was made between sociodemographic groups.

**Table 1:** Broad feature categories associated with PCD. Diagnosis, procedure, and drug codes were captured and characterized into broad feature categories.

| DIAGNOSIS [28] | PROCEDURES [28] | DRUGS [12] |
| --- | --- | --- |
| AIRWAY - Atelectasis (COUNT)<br>AIRWAY - Breathing Issues (COVERAGE)<br>AIRWAY - Bronchiectasis (FIRST)<br>AIRWAY - Bronchitis (COVERAGE)<br>AIRWAY - Cough (COVERAGE)<br>AIRWAY - Pneumonia (COUNT)<br>AIRWAY - Serious Pulmonology Events (COUNT)<br>AIRWAY - Upper Airway Infections (COVERAGE)<br>CM - Congenital Malformations of Heart (PRESENCE)<br>CM - Congenital Malformations of Spleen (PRESENCE)<br>CM - Congenital Malformations of Other Organs (PRESENCE)<br>CM - Unspecified Congenital Malformations (PRESENCE)<br>CM - Situs Inversus (PRESENCE)<br>DIAG. - Asthma (COUNT)<br>DIAG. - COVID-19 (COUNT)<br>DIAG. - Cystic Fibrosis (COUNT)<br>DIAG. - Pulmonary Diagnoses (COUNT)<br>DIAG. - Q348 - Congenital Malf. Resp. Sys. (Specific) (PRESENCE)<br>EAR - Hearing Loss (COUNT)<br>EAR - Otitis media (COVERAGE)<br>EAR - Otorrhea (COVERAGE)<br>FERTILITY - Infertility (COVERAGE)<br>IMMUNE SYSTEM - Immunodeficiency (COVERAGE)<br>NOSE - Nasal Congestion (COVERAGE)<br>NOSE - Nasal Polyps (COVERAGE)<br>NOSE - Rhinitis (COVERAGE)<br>QOL - Depression or Anxiety (COVERAGE)<br>SINUS - Chronic Sinusitis (COVERAGE) | PROC. - Acapella/Flutter: (COUNT)<br>PROC. - Adenoidectomy: (COUNT)<br>PROC. - BAHA: (COUNT)<br>PROC. - Bronchoalveolar lavage: (COVERAGE)<br>PROC. - Airway Clearance: (COVERAGE)<br>PROC. - Chest X-Rays: (COVERAGE)<br>PROC. - Cochlear Implant Maintenance: (COVERAGE)<br>PROC. - Cochlear Implant Placement: (COUNT)<br>PROC. - Detection of Microorganisms: (COVERAGE)<br>PROC. - Diagnostic EM: (PRESENCE)<br>PROC. - Endoscopic airway exam: (COVERAGE)<br>PROC. - Genetic Testing: (COUNT)<br>PROC. - Hearing aid Fitting + Maintenance: (COVERAGE)<br>PROC. - Hospitalization: (COUNT)<br>PROC. - IPV: (COUNT)<br>PROC. - Lung Function Measurement: (COVERAGE)<br>PROC. - Lung Transplant: (COUNT)<br>PROC. - Mastoidectomy: (COUNT)<br>PROC. - Myringotomy Tubes: (COUNT)<br>PROC. - Nasal Biopsy: (COUNT)<br>PROC. - Nitric Oxide Measurement: (COVERAGE)<br>PROC. - Oscillatory positive expiratory pressure: (COUNT)<br>PROC. - Sinus Surgery: (COVERAGE)<br>PROC. - Sputum Induction: (COVERAGE)<br>PROC. - Steroid Injection: (COVERAGE)<br>PROC. - Sweat Chloride Test: (COUNT)<br>PROC. - Tonsil surgery: (COUNT)<br>PROC. - Tympanoplasty: (COUNT) | DRUG - Anti-inflammatory (Non-Steroid)<br>DRUG - Anti-inflammatory (Steroid)<br>DRUG - Antibiotics<br>DRUG - Antidepressant<br>DRUG - Antifungals<br>DRUG - Antihistamine<br>DRUG - Bronchodilators<br>DRUG - Decongestant<br>DRUG - Expectorant<br>DRUG - Inhaled Hypertonic Saline<br>DRUG - Mucolytics<br>DRUG - Prophylactic Azithromycin |

### Training Cohorts and Datasets

The PCDF received IRB approval to use registry data. We identified 152 patients in the PCDFR who were definitively diagnosed with PCD based on two pathogenic variants in a PCD-associated gene and/or an ultrastructural defect on ciliary transmission electron microscopy. Clinical data for these patients were collected across 36 North American PCDF specialty centers using standardized data entry fields.

To link registry and claims data while maintaining privacy, we employed ‘tokenization’ as the process by which individual patients are assigned a unique ID based on heuristic matching of personally-identifiable information within a secure environment so that de-identified tokens are used to match individual patients. All work on these deidentified tokens was performed in a ‘clean-room’ environment from which only aggregated patient data could be exported. This avoided the risk of leaking potentially identifiable information such as precise diagnostic codes matched to dates but still permitted the training and execution of ML models. To protect patient privacy, insurance claim data had all codes for neonatal events removed, as that could reveal birth dates accurately. We filtered for cases with payer-based (closed) claims, ensuring comprehensive capture of patient encounters. As the cohort was predominantly pediatric, downstream analyses were restricted to patients ≤18 years old. This defined the PCDFR sub-cohort, forming a gold-standard set of 82 PCD cases within the broader claims dataset (**Figure 1**).

To address data imbalance and the small size of the PCDFR sub-cohort, we created a second sub-cohort of pediatric patients with claims codes suggestive of PCD. These were defined as patients with a diagnosis code of Q34.8 (Specific Congenital Malformation of the Respiratory System) recorded within six months of undergoing an Electron Microscopy (EM) procedure (Q34.8+EM). EM is often performed when there is high clinical suspicion of PCD. Q34.8+EM codes are commonly seen in PCD cohorts.

We randomly sampled patients from the screening cohort matched to confirmed PCD cases by age, gender, and ZIP-3 codes. This provided a background class assumed to be PCD-negative despite originating from the screening cohort (**Figure 1**).

### Machine-learning

Claims data from the Komodo Healthcare Map were restricted to closed systems, ensuring complete encounter records. We developed numerical scoring mechanisms for claims data to act as ‘features’ for ML classifiers of patient records over a year of coverage **(Table 1)**.

This is a standard approach to classifying tabular data. If a diagnostic/procedural code represented a permanent condition, its presence in a given year was scored as 1.0, and 0.0 otherwise (“PRESENCE”). Codes indicating serious clinical events (e.g., surgeries or severe pulmonary incidents) were scored based on their count (“COUNT”). For chronic or common conditions, scores reflected the number of two-week periods in which codes appeared, capturing their persistent nature (“COVERAGE”). For drug prescriptions, scores represented the total number of days a specific drug type was prescribed annually. Features were computed for each year, using the highest annual score across all available years. Feature values were scaled based on the proportion of closed coverage in a given year. For example, a “COUNT” feature observed over six months was doubled to represent a 12-month period. ZIP-3 codes and birth year were included as numerical features.

These features were used in machine-learning analyses with SciKitLearn implementations. Random Forest models were trained on numerical scores and composite codes from confirmed PCD cases, alongside matched background patients presumed negative for PCD. An adaptive synthetic (ADASYN) sampling approach was applied to rebalance training data by generating synthetic positive examples.^17^ This involves using algorithms to create artificial data points that mimic the characteristics of real positive cases based on patterns learned from existing data. These synthetic examples are not actual patient records but are computationally generated to provide additional training data for machine learning models, particularly in situations where real positive case data is scarce. To ensure the models were trained on features independent of the outcome definition, the codes used to define the Q34.8+EM sub-cohort were excluded from models trained on this expanded positive class.

We ran 5-fold cross validation classification experiments to assess the positive predictive value (precision) and sensitivity (recall) of each model.^18^. 5-fold cross-validation is a technique to evaluate a machine learning model by dividing the data into five parts, training and testing the model five times with a different part held out each time. The average performance across these tests provides a more reliable estimate of how well the model will perform on new, unseen data and helps in model selection and hyperparameter tuning. It’s particularly useful when dealing with limited datasets. We also assessed feature importance, assigning a score to each input feature in the model, indicating its relative contribution to the model’s predictive power. A higher decrease in impurity generally means the feature has a larger impact on the model’s predictions.

The model predicted a positive PCD diagnosis based on inclusion in either the PCDF cohort or Q34.8+EM sub-cohort. We made no assessment of inclusion bias based on socioeconomic groups for inclusion of individuals in each group (which could have been influenced by access to disease foundation resources or medical insurance). Outcome assessment was categorical and not based on subjective interpretation. Predictions from trained models were compared to published prevalence estimates^3^ to assess alignment between the number of pediatric PCD cases anticipated and those identified within the screening cohort.

## RESULTS

### Approximation of the Number of Patients with PCD in the PCD Screening Cohort

We estimated the number of PCD cases a robust ML model would predict within the screening cohort in the claims database. With approximately 72.5 million children in the U.S. in 2022 and a PCD prevalence of 1 in 7,554, an estimated 9,598 children have PCD (**Figure 2A**). Next we estimated the number of these children we would expect to find within the claims database, based on the linkage rate of pediatric cases from the PCDFR. About 90.3% of confirmed pediatric PCD cases in the PCDFR were linked to the claims database. Assuming this linkage rate applies nationwide, approximately 8,667 pediatric PCD cases would be represented in the database (**Figure 2B**). A well-performing model would predict a similar number of cases within the limited screening cohort of individuals with claims features often observed in PCD (**Appendix-1**), with an estimated prevalence of 1 in 159 among 1.38 million pediatric patients in the limited screening cohort (**Figure 2C**).

**Figure 2.**
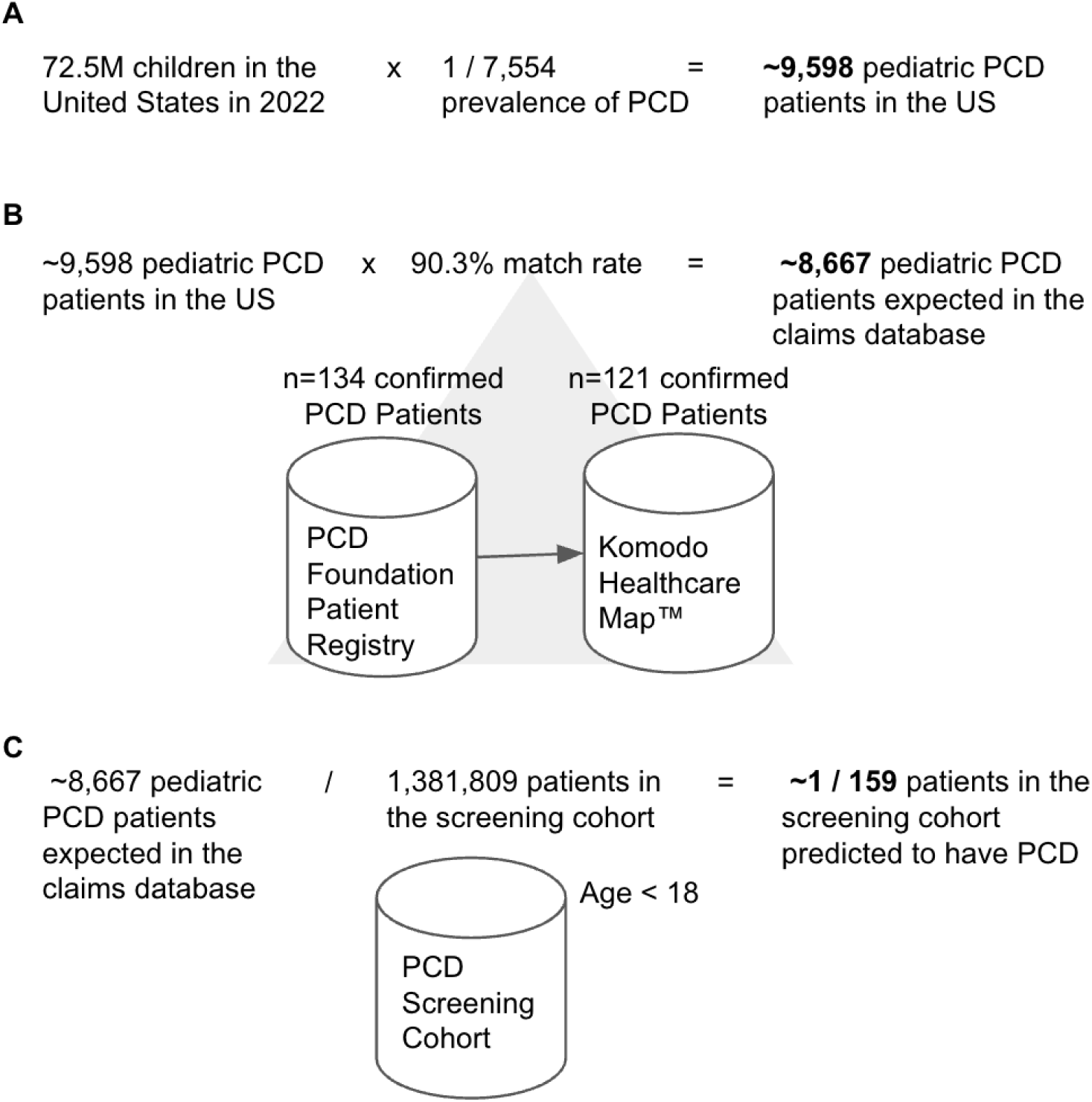
Estimation of patient counts in the claims database and prevalence within the screening cohort. **(A)** Rough calculation of the number of children in the United States who are living with PCD. (B) Approximation of the number of children with PCD who are captured within the claims database based on linkage rates of patients from the PCDFR. 121 pediatric patients from the PCDFR were successfully linked to cases in the claims database via tokenization. (C) Estimation of the prevalence of PCD within the screening cohort of ∼1.38M pediatric patients with claims features associated with PCD (Appendix-1).

Of the 136 patients between the claims database and PCDFR, downstream analysis focused on 82 confirmed pediatric PCD cases with closed claims, forming the PCDFR sub-cohort of the machine-learning positive class. An additional sub-cohort included 319 pediatric patients diagnosed with Q34.8 (Specific Congenital Malformation of the Respiratory System) within six months of an Electron Microscopy (EM) diagnostic procedure. These Q34.8+EM codes are frequently associated with PCD when there is high clinical suspicion.

### The PCD Screening Cohort Includes Patients With Clinical Features Associated With PCD And Related Conditions

We next analyzed claims codes across the screening cohort (**e-Appendix 1**) to characterize the population and the frequency of features often associated with PCD and related conditions. Analysis revealed the most frequent ICD codes were for congenital malformations, including 214,603 of 377,256 cases involving congenital heart defects (**Table 2**). Codes for cystic fibrosis (2.2%), bronchiectasis (2.6%), and infertility (1.7%) were less common. Some cases with PCD include cystic fibrosis codes, possibly due to misdiagnosis or coding limitations. We also identified combinations of clinical features linked to PCD, excluding cystic fibrosis. Examples include congenital malformations with bronchiectasis (0.28%), Q34.8 with situs inversus and bronchiectasis (0.02%), and situs inversus with bronchiectasis (0.03%) (**Table 2**). These findings indicate the screening cohort captures pediatric patients with PCD-like characteristics.

**Table 2.** Characteristics of the PCD screening cohort. Counts (and percentages) of patients within the PCD screening cohort of 1,381,809 pediatric patients with insurance claims codes associated with common PCD symptoms or clinical features, related conditions, or combinations of symptoms that could be considered to be patients at risk for PCD.

| Symptom or Clinical Feature | Patient Count |
| --- | --- |
| All congenital malformations | 377,256<br>(27.3%) |
| Congenital Malformations of Heart | 214,603<br>(15.5%) |
| Other Congenital Malformations | 210,937<br>(15.3%) |
| Situs Inversus | 9,799<br>(0.71%) |
| Cystic Fibrosis | 30,789<br>(2.2%) |
| Bronchiectasis | 36,307<br>(2.6%) |
| Infertility, >=12 years | 23,444<br>(1.7%) |
| Combination of all Congenital Malformations with Bronchiectasis, but without cystic fibrosis | 3,850<br>(0.28%) |
| Q34.8 with Situs Inversus and Bronchiectasis, but without cystic fibrosis, ≤12 years old | 238 (0.02%) |
| Situs Inversus with Bronchiectasis, but without cystic fibrosis | 370 (0.03%) |

### Performance evaluation supports the feasibility of developing machine-learning screening tools to identify patients likely to have PCD

We then trained random forest models to predict which patients in the screening cohort have PCD. We ran 5-fold cross validation classification experiments to assess the positive predictive value and sensitivity of each model. Using only PCDFR patients in comparison to background patients who were assumed to be PCD-negative, positive predictive value and sensitivity scores across different runs of the cross-validation process were highly variable and suggest that the small size of the PCDFR training set is problematic (**Figure 3A**). Augmentation did not improve model performance (**Figure 3B**).

**Figure 3.**
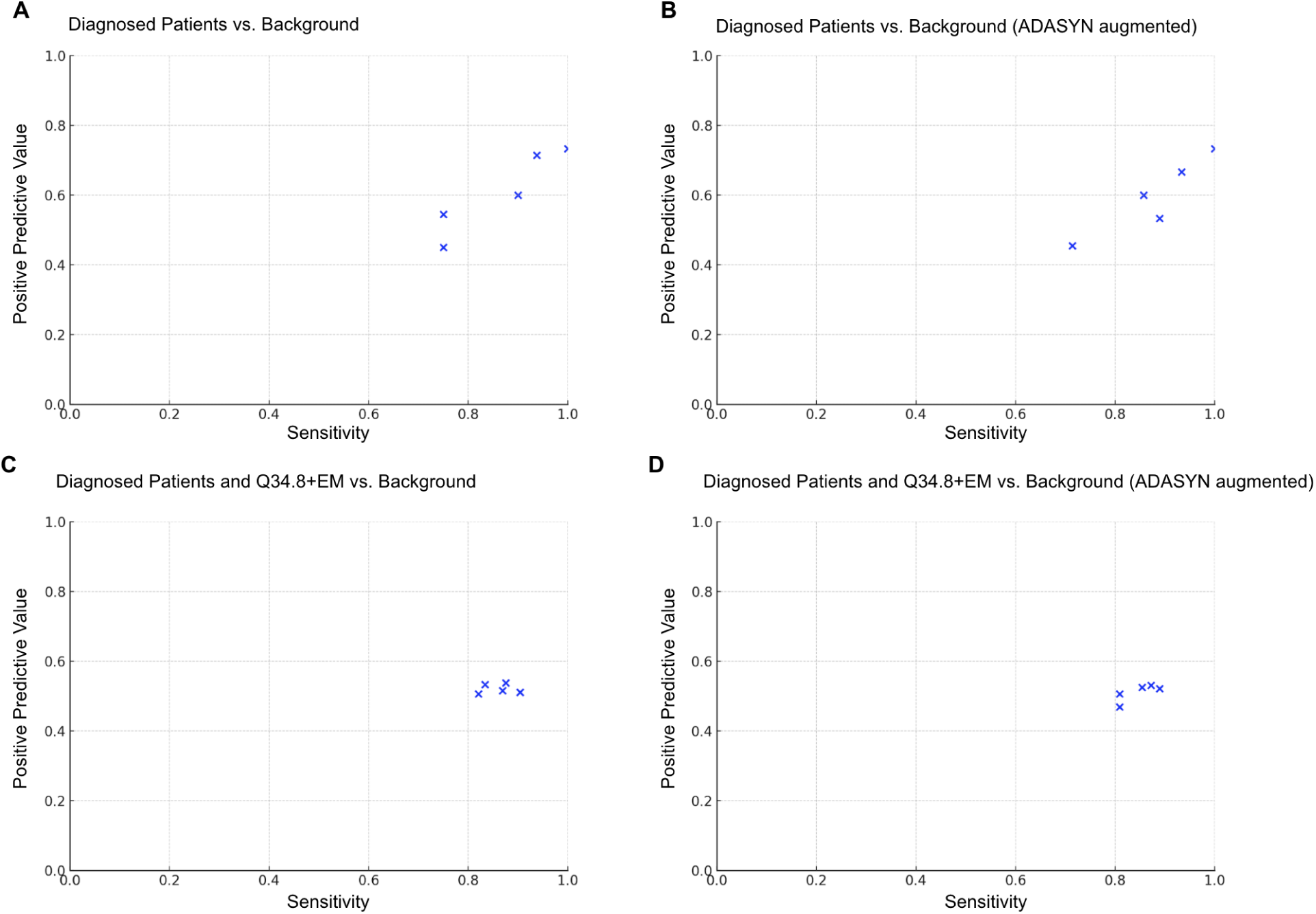
5-fold cross validation experiments on various training sets compared to randomly sampled background patients matched by age, gender, and zip3 codes. Positive predictive value-sensitivity plots with each metric reaching a maximum value of 1.0,. These metrics were generated from 5-fold cross validation experiments indicating perfect performance in identifying true positive cases without false positives or false negatives. Experiments compared randomly sampled background patients matched by age, gender, and zip3 codes to training sets including (A) diagnosed patients with PCD from the PCDFR (B) diagnosed patients with PCD from the PCDFR, ADASYN augmented, (C) diagnosed patients with PCD from the PCDFR plus patients with Q34.8 and EM claims codes, and (D) diagnosed patients with PCD from the PCDFR plus patients with Q34.8 and EM claims codes, ADASYN augmented.

Next we ran a 5-fold classification experiment using a training set of PCDFR patients combined with Q34.8+EM patients against background patients assumed to be PCD-negative. Positive predictive value and sensitivity scores were more tightly grouped, suggesting improved model performance with the inclusion of Q34.8+EM patients (**Figure 3C**). ADASYN augmentation did not further improve model performance (**Figure 3D**). We further applied this trained model to the entire database of 1,381,809 million patients who were aged ≤18 years and found that 7,705 patients were classified by the system as positive, within the order of magnitude of the expected 8,667 PCD patients (**Figure 2**). While we estimated a prevalence of 1 in 159, the model predicted that 1 in 179 patients in the screening cohort has PCD.

We then evaluated feature importance within the model predictions to determine the significance of each broad feature category included in the model (**e-Figure 1**). Among the 67 features used to train the model, the top 10 broad feature categories led to a 46.7% decrease in impurity (**Figure 4**). Three features (PROC. - BAHA, PROC. - Lung Transplant, and PROC. - Tympanoplasty) did not cause a decrease in impurity.

**Figure 4.**
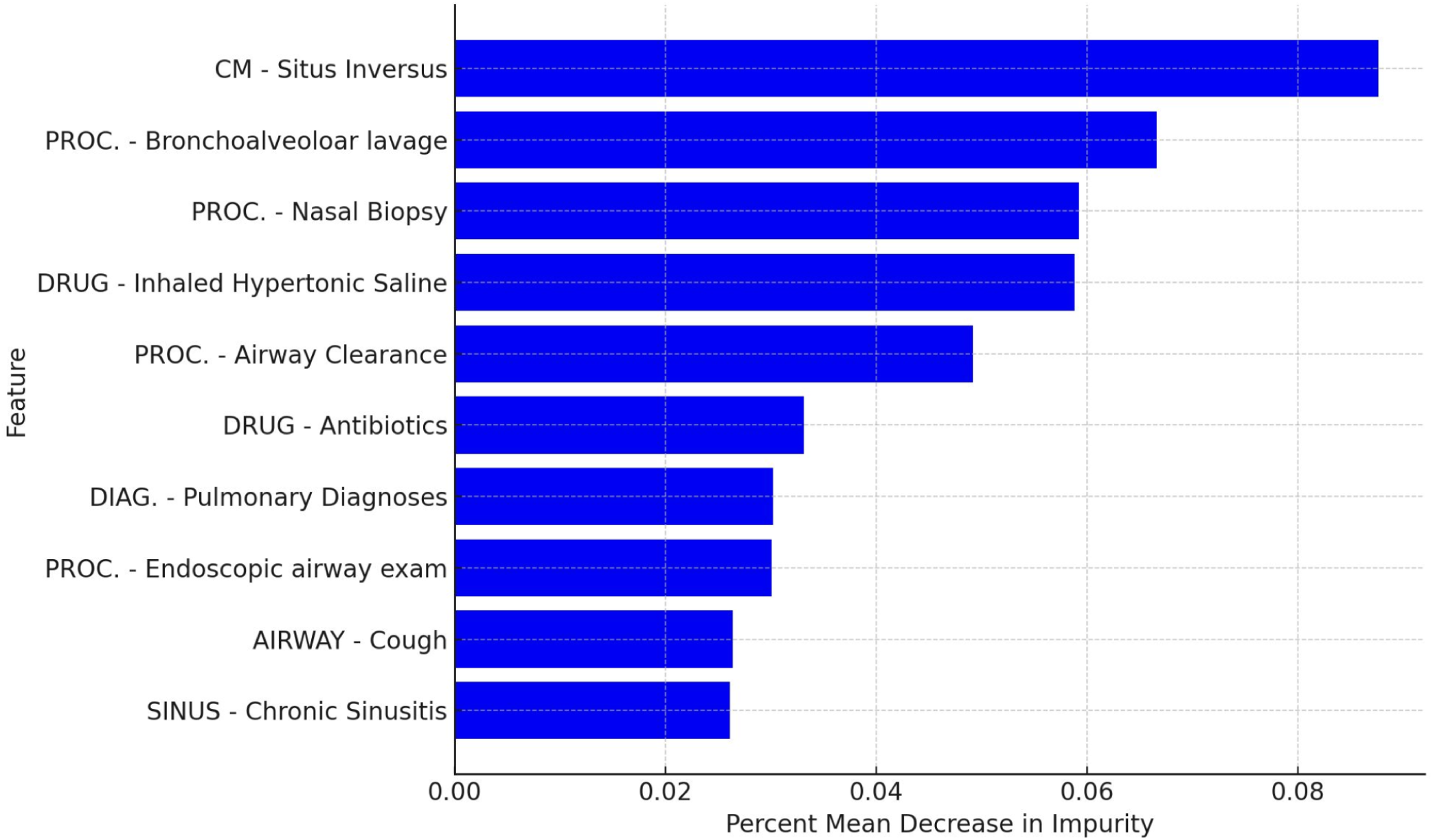
Plot of feature importance for the top 10 features based on mean decrease in impurity. Relative importance of the top 10 features determined by the mean decrease in impurity method using SciKitLearn. Percent mean decrease in impurity was calculated for each input feature in the model, indicating its relative contribution to the model’s predictive power. A higher decrease in impurity generally means the feature has a larger impact on the model’s predictions.

## DISCUSSION

Diagnostic delay remains a critical barrier impacting the lives of those living with PCD. We demonstrated the feasibility of integrating confirmed patient records from disease registries into large health insurance databases with national-level coverage, enabling the development of ML systems to identify individuals at high risk for PCD (as an example of a screening tool for rare disease). This effort was made possible through a multidisciplinary effort led by patient advocates, researchers, and clinicians to develop a detailed knowledge engineered representation of the insurance claims profile of patients with PCD. While unvalidated, this work may serve as the basis for future ML efforts in rare disease detection.

Although we did not validate our model on an independent dataset, we evaluated its performance using several key metrics. We assessed positive predictive value and sensitivity within a 5-fold cross-validation framework to measure how well the random forest model generalizes to unseen data. Notably, the inclusion of patients with Q34.8 and EM codes led to improved model performance (**Figure 3C**), suggesting that expanding the positive case pool can help mitigate the challenges of imbalanced datasets. This can be achieved through collaborations with patient organizations, medical centers, or potentially through AI-driven approaches, such as generating synthetic positive cases. Unexpectedly, ADASYN augmentation did not improve model performance in our tests, regardless of the threshold or training set composition, possibly due to the small size of the training data and complexity of the feature space (**Figure 3B,3D)**.

Another way to assess model performance is to compare the number of patients predicted to have PCD by the model with the expected number of cases in the screening cohort. The model classified 7,705 patients as positive, which aligns with 8,667 patients with PCD we anticipated based on our initial calculations (**Figure 2**). This result is promising; however, we cannot determine the true positive predictive value through actual PCD diagnostic testing since these patients remain deidentified.

We reviewed the relative importance of clinical features in the final model (**Figure 3D**). Initially, features were treated without predefined weighting. Features selected more frequently and contributing more to impurity reduction were assigned higher importance scores (**Figure 4**). Reducing impurity improves the model’s ability to correctly classify patients based on their PCD likelihood. The top feature was situs inversus, which led to an 8.76% reduction in impurity, compared to the average reduction of 1.56% across all features. This result is unsurprising, as slightly less than 50% of patients with PCD have situs inversus totalis, and an additional 12% have more complex laterality defects with *situs ambiguus*^19^. Several features that we might expect to see contributing to positive predictions were not among the top, such as airway clearance or inhaled hypertonic saline, which we speculate may have resulted from manual grouping of codes to form the broad feature categories. For example, we may have instead grouped “inhaled hypertonic saline” with “DRUG-mucolytics” as a broad feature category. Modern deep learning models have the advantage of automatically inferring optimal feature sets and thus would avoid this issue^20^.

### Limitations

There are several important limitations to this approach. First is the small size of our training set expressed as a set of positive cases linked from the PCDFR. We utilized hashing technology to identify confirmed cases from the PCDFR in the claims database, which increases the overall accuracy of identifying patients but also incurs an increased risk of ‘collisions’ where separate patients are incorrectly conflated into a single patient record. We also assumed that the background cohort in the claims database was PCD-negative. Further, in accordance with patient privacy protections, we were unable to reidentify and confirm the presence or absence of a PCD diagnosis in the Q34.8+EM sub-cohort of the positive class. This lack of individual-level confirmation introduces a limitation, as it is possible that EM was conducted due to suggestive history but ultimately yielded negative results, a scenario not captured in our population-level data.

The claims database on which we developed the training and screening cohorts was a unified, national-scale insurance claim database that provided widespread coverage to include many relevant populations, but did not capture neonatal populations. Given that situs inversus totalis with neonatal respiratory distress is sensitive and specific for PCD, future machine-learning methods should aim to include neonates in the study population. Medicare patients are not represented in the claims database. The inclusion of these populations may improve model performance in a general population^21^. There are also limitations inherent to the use of claims data. Notably, the presence of a procedure code does not guarantee a specific outcome or result, and the reporting of a drug code does not confirm if the prescription was actually filled and adhered to by the patient. Furthermore, claims data may lack granular clinical details, temporal information beyond service dates, and insights into patient behavior or lifestyle factors that could influence health outcomes.

This work serves as a foundational methodology, designed with a lightweight implementation to ensure it operates efficiently on a small-scale analysis platform. We used a tabulated approach that resulted in a reduced set of features that summarized patients’ clinical experiences in any given year and then used only the maximum feature value across all available years as the final feature used in the analysis.

### Future Directions

We demonstrated the feasibility of ML methods for patient screening for PCD based on national-level insurance claim data in the absence of an ICD code^22^. While the approach used closed claims data and manual feature categorization for a random forest model, future ML models could leverage more powerful algorithms, trained on hundreds of features, including time-series data, to further improve classification accuracy. Future efforts could explore the use of national electronic medical record data to train neural networks, as this data more accurately reflects the clinical environments in which such screening tools will be applied^23,24^.

The key challenge when developing ML-based tools for rare diseases is the relatively small number of available patients. Patient-led organizations are making rapid strides towards the development and utilization of research-ready, rare disease patient data for natural history and clinical studies. The PCDF is one such example, establishing a clinical registry in 2020 to collect rigorous and detailed diagnostic and phenotypic data on individuals with genetically confirmed PCD through the PCDF Clinical and Research Centers Network, and expanding the PCDFR from approximately 150 patient participants at the time of linkage and analysis in this study, to now over 600 participants from 36 North American specialty centers accredited in diagnosis and management of patients with PCD. These efforts are providing crucial infrastructure to drive research partnerships and will be instrumental in the pursuit of improved screening, diagnosis, and care for the PCD community.

As patient organizations and their partners continue to develop comprehensive registries and datasets, there is a profound opportunity to scale ML-based approaches for screening many of the estimated 300 million people worldwide living with rare diseases. Once validated, these tools could be deployed in diverse clinical settings, including in international communities with significant disparities in access to diagnosis and care^25^, enabling rapid identification of patients for referral and significantly reducing the time from first clinic visit to diagnostic testing. ML has the potential to transform the diagnostic landscape, bringing timely and accurate diagnoses to those who have long faced a complex diagnostic journey.

## CONCLUSIONS

Our study is the first to demonstrate the feasibility of ML for PCD screening using national insurance claim data. Initial model performance was suboptimal, but improvements were made by expanding the positive class dataset. Future work may develop and validate neural network models for PCD, aiding in the identification of patients who may benefit from timely diagnostic testing and targeted interventions.

## Data Availability

All data produced in the present study are available upon reasonable request to the authors

## ABBREVIATION LIST

PCD: primary ciliary dyskinesia
Q34.8+EM: presence of an Q34.8 diagnosis code within six months of an electron microscopy procedure code
ML: machine learning
PCDFR: PCD Foundation Registry
ICD: International Classification of Disease
ADASYN: adaptive synthetic

## DECLARATIONS

### Ethics approval and consent to participate

<u>Institutional Review Board</u>: Protocol number PCDFR001 approved by the Genetic Alliance Institutional Review Board. This study was not registered.

### Consent for publication

Not applicable

### Availability of data and materials

We will include source code of our methods as an open source library available for any future users of the Komodo’s Healthcare Map.

### Authors’ contributions

Conceptualization: G.B. C.K. M.M. C.M. M.G.O, A.J.S, H.B-P

Data Curation: G.B. C.K. M.M. C.M. R.P. M.G.O, A.J.S, H.B-P

Formal Analysis: G.B. C.K. M.M. C.M. R.P. M.G.O, A.J.S, H.B-P

Methodology: G.B. C.K. M.M. C.M. R.P. M.G.O A.J.S H.B-P

Project Administration: G.B. R.P. H.B-P Software: G.B.

Visualization: G.B.

Writing – Original Draft Preparation: G.B. C.K. M.M. C.M. R.P. M.G.O, A.J.S, H.B-P

Writing – Review & Editing: G.B. C.K. M.M. C.M. R.P. M.G.O, A.J.S, H.B-P

## Acknowledgements

This work was supported by the Chan Zuckerberg Initiative Foundation’s Rare as One Program, as well as generous accommodation for performing this work from both Komodo and Datavant. Thanks to the PCD Foundation for work developing a clinical-grade registry and an IRB application to the Genetic Alliance, and for their leadership and vision in developing this work. Thanks to Juliane Mills for her contributions to the framing of the project. Thank you to the patients with PCD who have participated in the PCDFR for sharing your data and enabling this work.

## Authors’ information (optional)

Not applicable

---

**e-Figure 1.**
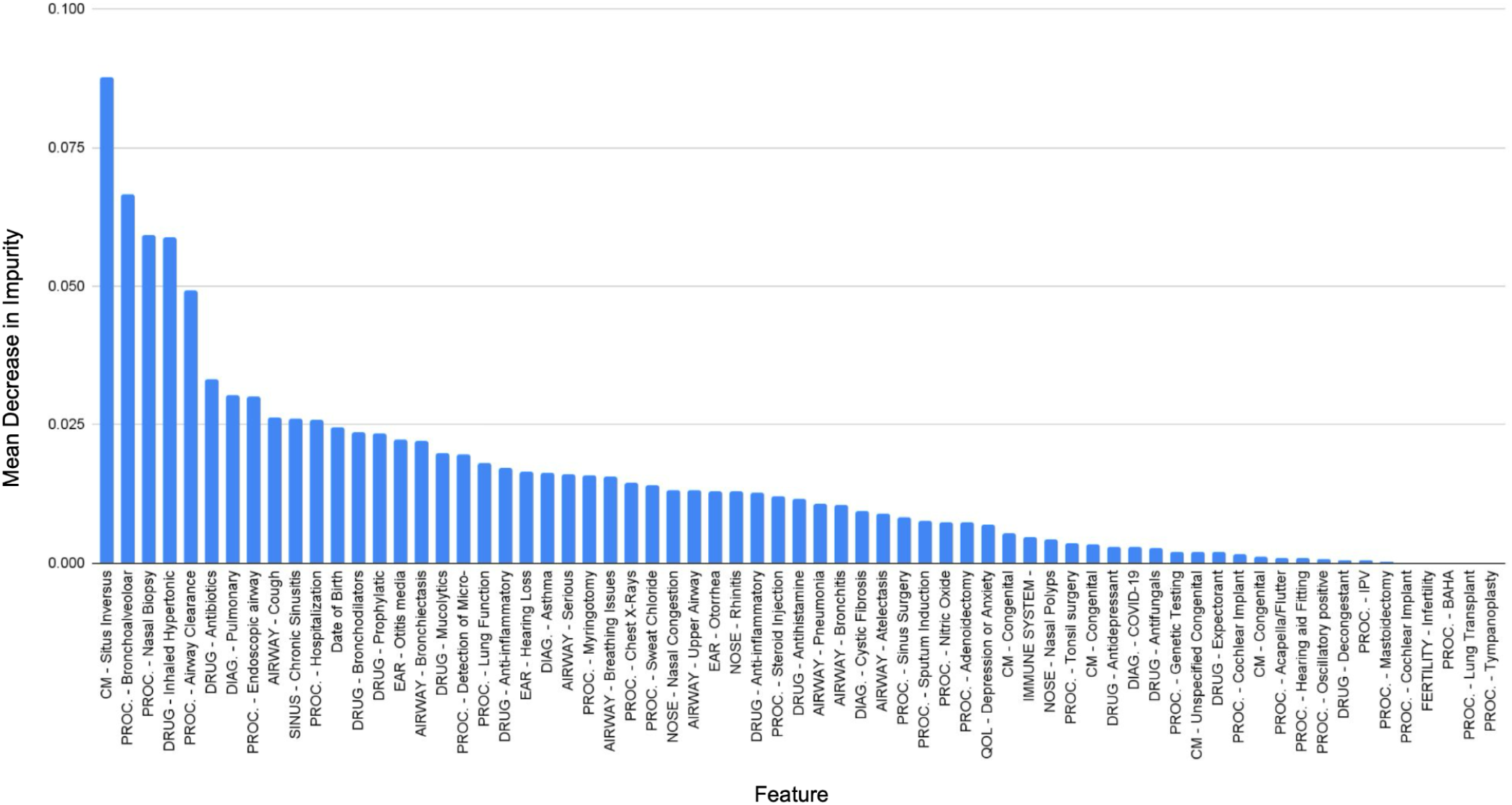
Plot of feature importance for features based on mean decrease in impurity. Relative importance of the all included features determined by the mean decrease in impurity method using SciKitLearn. Vertical bars represent the importance scores, with higher bars indicating greater importance within the random forest model.

