## Supplementary material for "Feasibility of Machine Learning Analysis for the Identification of Patients with Possible Primary Ciliary Dyskinesia": e-Appendix-1

### e-Appendix-1: Definition of the Initial PCD Cohort with the Komodo Healthcare Map

Table of Contents

Introduction 1

ICD Codes 1

1. All ‘Standard’ General Codes for PCD 1
2. Codes for Common Conditions that are generally more frequent and regular in PCD for children aged <12 1
3. Catch-all Code for adults: Bronchiectasis OR Infertility 4
4. Cystic Fibrosis 4
5. Congenital Malformations (Situs Inversus, Heterotaxy, etc) 4
6. ICU for infants + cough / upper respiratory / nasal congestion 5

Query Structure + Output 5

#### INTRODUCTION

This document describes the initial query upon which all subsequent analysis will be performed in this study. We seek to provide a comprehensive query on Komodo Health’s Prism system that will cover all patients with PCD and a sizable background cohort that will include patients that are correctly diagnosed with the codes that patients with PCD are often misdiagnosed for. The challenge is that the PCD symptoms involve very common infections and conditions that impact large numbers of people.

We resolve this by structuring our query as a composite ‘OR’ made up of complex subqueries. We seek here to confirm that the codes (and logic) used in these subqueries are accurate and correct and that we are not missing important clinically relevant codes.

#### ICD CODES

##### 1. All ‘Standard’ General Codes for PCD

- Q348 - Other Specified Congenital Malformations Of Respiratory System (8.6k)
- Q349 - Congenital Malformation Of Respiratory System, Unspecified (23k)
- Q34 - Other Congenital Malformations Of Respiratory System (1.2k)

##### 2. Codes for Common Conditions that are generally more frequent and regular in PCD for children aged <12

Search for 9 or more visits for *any* of the following codes over periods of 18 months from 3/1/2105 - 8/31/2022 (i.e., approximately 1 visit every 2 months)

- Chronic Otitis Media:

- 3813 - Other And Unspecified Chronic Nonsuppurative Otitis Media (186k)
- 3821 - Chronic Tubotympanic Suppurative Otitis Media (30k)
- 3822 - Chronic Atticoantral Suppurative Otitis Media (42k)
- 3823 - Unspecified Chronic Suppurative Otitis Media (110k)
- 38110 - Chronic Serous Otitis Media, Simple Or Unspecified (474k)
- 38119 - Other Chronic Serous Otitis Media (82k)
- 38120 - Chronic Mucoid Otitis Media, Simple Or Unspecified (208k)
- 38129 - Other Chronic Mucoid Otitis Media (19k)
- H663X1 - Other Chronic Suppurative Otitis Media, Right Ear (62k)
- H663X2 - Other Chronic Suppurative Otitis Media, Left Ear (59k)
- H663X3 - Other Chronic Suppurative Otitis Media, Bilateral (159k)
- H663X9 - Other Chronic Suppurative Otitis Media, Unspecified Ear (43k)
- H6520 - Chronic Serous Otitis Media, Unspecified Ear (237k)
- H6521 - Chronic Serous Otitis Media, Right Ear (336k)
- H6522 - Chronic Serous Otitis Media, Left Ear (316k)
- H6523 - Chronic Serous Otitis Media, Bilateral (1.2m)
- H6530 - Chronic Mucoid Otitis Media, Unspecified Ear (52k)
- H6531 - Chronic Mucoid Otitis Media, Right Ear (88k)
- H6532 - Chronic Mucoid Otitis Media, Left Ear (83k)
- H6533 - Chronic Mucoid Otitis Media, Bilateral (423k)
- H6611 - Chronic Tubotympanic Suppurative Otitis Media, Right Ear (33k)
- H6612 - Chronic Tubotympanic Suppurative Otitis Media, Left Ear (31k)
- H6613 - Chronic Tubotympanic Suppurative Otitis Media, Bilateral (84k)
- H6623 - Chronic Atticoantral Suppurative Otitis Media, Bilateral (24k)
- H65413 - Chronic Allergic Otitis Media, Bilateral (19k)
- H65491 - Other Chronic Nonsuppurative Otitis Media, Right Ear (72k)
- H65492 - Other Chronic Nonsuppurative Otitis Media, Left Ear (68k)
- H65493 - Other Chronic Nonsuppurative Otitis Media, Bilateral (404k)
- H65499 - Other Chronic Nonsuppurative Otitis Media, Unspecified Ear (85k)

- Upper Airway Infections:
  - J069 - Acute Upper Respiratory Infection, Unspecified (66m)
  - 4659 - Acute Upper Respiratory Infections Of Unspecified Site (34m)
  - 4658 - Acute Upper Respiratory Infections Of Other Multiple Sites (1.7m)
- Pneumonia: *[Searched for Pneumonia - excluded codes that specified 'viral']*
  - J189 - Pneumonia, Unspecified Organism (18m)
  - 486 - Pneumonia, Organism Unspecified (9.2m)
  - J181 - Lobar Pneumonia, Unspecified Organism (3.9m)
  - J159 - Unspecified Bacterial Pneumonia (1.8m)
  - Z8701 - Personal History Of Pneumonia (Recurrent) (1.8m)
  - J188 - Other Pneumonia, Unspecified Organism (858k)
  - 481 - Pneumococcal Pneumonia [Streptococcus Pneumoniae Pneumonia] (275k)
  - J157 - Pneumonia Due To Mycoplasma Pneumoniae (342k)
  - 4829 - Bacterial Pneumonia, Unspecified (376k)

    - J13 - Pneumonia Due To Streptococcus Pneumoniae (231k)
    - J168 - Pneumonia Due To Other Specified Infectious Organisms (507k)
    - J158 - Pneumonia Due To Other Specified Bacteria (344k)
    - V1261 - Personal History Of Pneumonia (Recurrent) (246k)
    - J150 - Pneumonia Due To Klebsiella Pneumoniae (135k)
    - J151 - Pneumonia Due To Pseudomonas (188k)
    - J156 - Pneumonia Due To Other Gram-Negative Bacteria (282k)
    - J15212 - Pneumonia Due To Methicillin Resistant Staphylococcus Aureus (182k)
    - 4838 - Pneumonia Due To Other Specified Organism (141k)
- Wet Cough:
    - R05 - Cough (76m)
    - 7862 - Cough (34m)
- Nasal Congestion:
    - R0981 - Nasal Congestion (18m)
- Atelectasis:
    - J9811 - Atelectasis (9.1m)
    - Other codes pertaining to neonates have been scrubbed
- Protracted Bacterial Bronchitis
    - J42 - Unspecified Chronic Bronchitis (1.4m)
- Chronic Bronchitis
    - 4910 - Simple Chronic Bronchitis (161k)
    - 4911 - Mucopurulent Chronic Bronchitis (95k)
    - 4918 - Other Chronic Bronchitis (90k)
    - 4919 - Unspecified Chronic Bronchitis (540k)
    - 49120 - Obstructive Chronic Bronchitis Without Exacerbation (1.2m)
    - 49121 - Obstructive Chronic Bronchitis With (Acute) Exacerbation (3.0m)
    - 49122 - Obstructive Chronic Bronchitis With Acute Bronchitis (276k)
    - J41 - Simple And Mucopurulent Chronic Bronchitis
    - J42 - Unspecified Chronic Bronchitis (1.4m)
    - J410 - Simple Chronic Bronchitis (942k)
    - J411 - Mucopurulent Chronic Bronchitis (292k)
    - J418 - Mixed Simple And Mucopurulent Chronic Bronchitis (154k)
- Chronic Sinusitis
    - J329 - Chronic Sinusitis, Unspecified (13m)
    - 4739 - Unspecified Sinusitis (Chronic) (9.5m)
    - J320 - Chronic Maxillary Sinusitis (3.6m)
    - 4730 - Chronic Maxillary Sinusitis (1.4m)
    - J322 - Chronic Ethmoidal Sinusitis (1.4m)
    - J328 - Other Chronic Sinusitis (1.4m)
    - J321 - Chronic Frontal Sinusitis (1.1m)
    - 4738 - Other Chronic Sinusitis (742k)
    - J323 - Chronic Sphenoidal Sinusitis (593k)
    - 4732 - Chronic Ethmoidal Sinusitis (388k)
    - 4731 - Chronic Frontal Sinusitis (262k)

- 4733 - Chronic Sphenoidal Sinusitis (89k)
    - J32 - Chronic Sinusitis (3.6k)
- Severe Asthma
    - J45991 - Cough Variant Asthma (1.1m)
    - J4550 - Severe Persistent Asthma, Uncomplicated (636k)
    - J4551 - Severe Persistent Asthma With (Acute) Exacerbation (366k)
    - J4552 - Severe Persistent Asthma With Status Asthmaticus (72k)
    - J455 - Severe Persistent Asthma (790)
- Tachypnea (suggested by Dr. Milla)
    - R0682 - Tachypnea, Not Elsewhere Classified (1.0m)
    - 78606 - Tachypnea (352k)

**3. Catch-all Code for adults: Bronchiectasis OR Infertility**

- Bronchiectasis
    - J479 - Bronchiectasis, Uncomplicated (1.1m)
    - 4940 - Bronchiectasis Without Acute Exacerbation (202k)
    - J471 - Bronchiectasis With (Acute) Exacerbation (192k)
    - J470 - Bronchiectasis With Acute Lower Respiratory Infection (137k)
    - 4941 - Bronchiectasis With Acute Exacerbation (105k)
    - Q334 - Congenital Bronchiectasis (3.5k)
    - 74861 - Congenital Bronchiectasis (1.8k)
    - J47 - Bronchiectasis (1.3k)
- Infertility
    - N979 - Female Infertility, Unspecified (1.2m)
    - N469 - Male Infertility, Unspecified (340k)
    - N978 - Female Infertility Of Other Origin (306k)
    - 6289 - Infertility, Female, Of Unspecified Origin (257k)
    - N971 - Female Infertility Of Tubal Origin (137k)
    - 6069 - Male Infertility, Unspecified (72k)
    - 6282 - Infertility, Female, Of Tubal Origin (71k)
    - 6288 - Infertility, Female, Of Other Specified Origin (50k)
    - N468 - Other Male Infertility (46k)
    - N97 - Female Infertility (4.2k)
    - N46 - Male Infertility (3.8k)

**4. Cystic Fibrosis, except nonpulmonary manifestations of CF.**

- E849 - Cystic Fibrosis, Unspecified (93k)
- E840 - Cystic Fibrosis With Pulmonary Manifestations (55k)
- 27702 - Cystic Fibrosis With Pulmonary Manifestations (17k)
- E84 - Cystic Fibrosis (1.5k)

**5. Congenital Malformations (Situs Inversus, Heterotaxy, etc)**

Including only internal organs, not brain or spinal cord.

- Q248 - Other Specified Congenital Malformations Of Heart (180k)

- Q892 - Congenital Malformations Of Other Endocrine Glands (146k)
- Q899 - Congenital Malformation, Unspecified (124k)
- Q898 - Other Specified Congenital Malformations (83k)
- Q897 - Multiple Congenital Malformations, Not Elsewhere Classified (83k)
- Q228 - Other Congenital Malformations Of Tricuspid Valve (73k)
- Q638 - Other Specified Congenital Malformations Of Kidney (66k)
- Q453 - Other Congenital Malformations Of Pancreas And Pancreatic Duct (69k)
- Q8901 - Asplenia (Congenital) (52k)
- Q238 - Other Congenital Malformations Of Aortic And Mitral Valves (43k)
- Q893 - Situs Inversus (30k)
- Q8909 - Congenital Malformations Of Spleen (25k)
- Q891 - Congenital Malformations Of Adrenal Gland (12k)
- Q89 - Other Congenital Malformations, Not Elsewhere Classified (3.5k)
- Q890 - Congenital Absence And Malformations Of Spleen (14)

**6. ICU for infants + cough / upper respiratory / nasal congestion**

- PROC:99477 + 0 ≤ age ≤ 2 + ( ICD:R05, ICD:J069, ICD:4659, ICD:R0981 between 0-7 days before event and 0-7 days after event)

#### QUERY STRUCTURE + OUTPUT

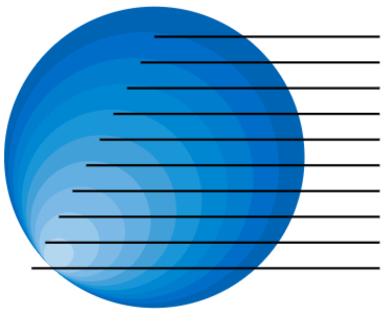

| | Patients | HCPs | HCOs |
|---|---|---|---|
| Patient Filter 1: 3/1/2021 - 8/30/2022 | 156,908 | 86,600 | 24,497 |
| Patient Filter 2: 9/1/2019 - 2/28/2021 | 293,214 | 132,873 | 34,702 |
| Patient Filter 3: 3/1/2018 - 8/30/2019 | 566,863 | 193,189 | 46,540 |
| Patient Filter 4: 9/1/2016 - 2/28/2018 | 878,423 | 238,291 | 56,701 |
| Patient Filter 5: 3/1/2015 - 8/30/2016 | 1,177,783 | 270,353 | 64,546 |
| Patient Filter 6: Standard PCD Codes | 1,211,928 | 281,103 | 66,379 |
| Patient Filter 7: Bronchiectasis / Inferti... | 5,732,950 | 756,454 | 161,986 |
| Patient Filter 8: Cystic Fibrosis | 5,842,132 | 794,489 | 168,776 |
| Patient Filter 9: Congenital Malformations... | 6,680,312 | 872,976 | 179,440 |
| Patient Filter 10: ICU for infants + cough ... | 6,687,486 | 873,145 | 179,449 |

Note that queries 1-5 refer to the section marked **'Codes for Common Conditions that are generally more frequent and regular in PCD for children aged <12'** and are time-delineated because they denote 9+ visits over all ICD conditions listed in the 18 month periods shown**.**
