## Supplementary material for "Feasibility of Machine Learning Analysis for the Identification of Patients with Possible Primary Ciliary Dyskinesia": e-Appendix-2

### e-Appendix-2: ICD10 / CPT / Drug Coding Reference for Primary Ciliary Dyskinesia


**INTRODUCTION** 3
**DIAGNOSIS CATEGORIES AND MEASURES** 3
**ICD DIAGNOSIS CODES** 5
AIRWAY - Atelectasis 5
AIRWAY - Breathing Issues 5
AIRWAY - Bronchiectasis 5
AIRWAY - Bronchitis 5
AIRWAY - Cough 6
AIRWAY - Serious Pulmonology Events 6
AIRWAY - Pneumonia 7
AIRWAY - Upper Airway Infections 7
CM - Congenital Malformations of Heart 7
CM - Other Congenital Malformations 8
CM - Situs Inversus 8
DIAG. - Asthma 8
DIAG. - COVID-19 8
DIAG. - Cystic Fibrosis 9
DIAG. - Pulmonary Diagnoses 9
DIAG. - Q348 - Congenital Malf. Resp. Sys. (Specific) 9
EAR - Otitis media 9
EAR - Hearing Loss 10
EAR - Otorrhea 12
FERTILITY - Infertility 12
IMMUNE SYSTEM - Immunodeficiency 12
NOSE - Nasal Congestion 13
NOSE - Nasal Polyps 13
NOSE - Rhinitis 13
QOL - Depression or Anxiety 13
SINUS - Chronic Sinusitis 14
**PROCEDURE CATEGORIES** 14
**PROCEDURE CODES** 15
PROC. - Acapella/Flutter 15
PROC. - Adenoidectomy 15
PROC. - BAHA 15
PROC. - Bronchoalveoloar lavage 15
PROC. - Cochlear Implant Placement 17
PROC. - Cochlear Implant Maintenance 18
PROC. - Chest Wall Manipulation 19
PROC. - Chest X-Rays 19
PROC. - Diagnostic EM 19
PROC. - Endoscopic airway exam 19

PROC. - Genetic Testing 19
PROC. - Hearing aid Fitting + Maintenance 20
PROC. - Hospitalization 22
PROC. - IPV 22
PROC. - Lung Function Measurement 22
PROC. - Lung Transplant 23
PROC. - Mastoidectomy 23
PROC. - Myringotomy Tubes 24
PROC. - Nasal Biopsy 24
PROC. - Nitric Oxide Measurement 24
PROC. - Oscillatory positive expiratory pressure 24
PROC. - Sinus Surgery 24
PROC. - Detection of Micro-organsms 26
PROC. - Sputum Induction 27
PROC. - Steroid Injection 27
PROC. - Sweat Chloride Test 27
PROC. - Tonsil surgery 27
PROC. - Tympanoplasty 28
**DRUG CODES 28**
DRUG - Antibiotics 29
DRUG - Antidepressant 30
DRUG - Antifungals 30
DRUG - Antihistamine 30
DRUG - Anti-inflammatory (Steroid) 30
DRUG - Anti-inflammatory (Non-Steroid) 30
DRUG - Bronchodilators 31
DRUG - Decongestant 31
DRUG - Expectorant 31
DRUG - Mucolytics 31

#### INTRODUCTION

This document describes how we are coding and capturing features for computational analysis with machine learning. At the current time this data is expressed as a list of codes grouped into categories. We have defined 28 categories for Diagnoses, 28 for Procedures, and 12 for Drug Prescriptions. These are fully described below.

#### DIAGNOSIS CATEGORIES AND MEASURES

We split the main categories into diagnoses pertaining to (A) issues of the airway and of breathing; (B) congenital malformations; (C) broad categories of diagnoses and possible misdiagnoses; (D) ear, nose, and sinus-related infections and issues; (E) fertility issues; (F) quality of life measures to characterize how the disease impacts patients experience more deeply.

Each category is assigned a feature type in parentheses and **COUNT**, **COVERAGE**, or **FIRST** in a way that determines how the feature is computed.

**COUNT** features are simply the number of separate days that a diagnosis in that category occurs in the course of a year. These groups of diagnosis codes are typically serious events (such as a collapsed lung, a pneumonia diagnosis, or the presence of COVID-19).

**COVERAGE** features are the number of separate two-week periods in which a diagnosis from that category appears over the course of a year. These events are thought to be more chronic and interconnected, so that we treat one occurrence over the two weeks the same as 5 similar events in the same time period.

**FIRST** features are binary features that are set to 0.0 before they first occur in the longitudinal data and 1.0 after that. This coding scheme denotes the presence of conditions that don't change over time (e.g,. Situs Inversus or other congenital malformations, bronchiectasis, etc).

The categories of diagnostic features being used are:

- AIRWAY - Atelectasis (COUNT)
- AIRWAY - Breathing Issues (COVERAGE)
- AIRWAY - Bronchiectasis (FIRST)
- AIRWAY - Bronchitis (COVERAGE)
- AIRWAY - Cough (COVERAGE)
- AIRWAY - Pneumonia (COUNT)
- AIRWAY - Serious Pulmonology Events (COUNT)
- AIRWAY - Upper Airway Infections (COVERAGE)
- CM - Congenital Malformations of Heart (FIRST)
- CM - Congenital Malformations of Spleen (FIRST)
- CM - Congenital Malformations of Other Organs (FIRST)
- CM - Unspecified Congenital Malformations (FIRST)
- CM - Situs Inversus (FIRST)
- DIAG. - Asthma (COUNT)
- DIAG. - COVID-19 (COUNT)
- DIAG. - Cystic Fibrosis (COUNT)
- DIAG. - Pulmonary Diagnoses (COUNT)
- DIAG. - Q348 - Congenital Malf. Resp. Sys. (Specific) (FIRST)
- EAR - Hearing Loss (COUNT)

- EAR - Otitis media (COVERAGE)
- EAR - Otorrhea (COVERAGE)
- FERTILITY - Infertility (COVERAGE)
- IMMUNE SYSTEM - Immunodeficiency (COVERAGE)
- NOSE - Nasal Congestion (COVERAGE)
- NOSE - Nasal Polyps (COVERAGE)
- NOSE - Rhinitis (COVERAGE)
- QOL - Depression or Anxiety (COVERAGE)
- SINUS - Chronic Sinusitis (COVERAGE)

**ICD DIAGNOSIS CODES**

AIRWAY - Atelectasis

J9811 - Atelectasis

AIRWAY - Breathing Issues

78606 - Tachypnea
R062 - Wheezing
R069 - Unspecified Abnormalities Of Breathing
R0600 - Dyspnea, Unspecified
R0602 - Shortness Of Breath
R0682 - Tachypnea, Not Elsewhere Classified
R0689 - Other Abnormalities Of Breathing

AIRWAY - Bronchiectasis

4940 - Bronchiectasis Without Acute Exacerbation
4941 - Bronchiectasis With Acute Exacerbation
74861 - Congenital Bronchiectasis (1.8k)
J47 - Bronchiectasis (1.3k)
J470 - Bronchiectasis With Acute Lower Respiratory Infection
J471 - Bronchiectasis With (Acute) Exacerbation
J479 - Bronchiectasis, Uncomplicated
Q334 - Congenital Bronchiectasis (3.5k)

AIRWAY - Bronchitis

466 - Acute bronchitis and bronchiolitis
490 - Bronchitis, not specified as acute or chronic
491 - Chronic bronchitis
4660 - Acute bronchitis
4910 - Simple chronic bronchitis
4910 - Simple Chronic Bronchitis
4911 - Mucopurulent Chronic Bronchitis
4912 - Obstructive chronic bronchitis
4918 - Other Chronic Bronchitis
4919 - Unspecified Chronic Bronchitis
5060 - Bronchitis and pneumonitis due to fumes and vapors
49120 - Obstructive Chronic Bronchitis Without Exacerbation
49121 - Obstructive Chronic Bronchitis With (Acute) Exacerbation
49122 - Obstructive Chronic Bronchitis With Acute Bronchitis
J20 - Acute bronchitis
J40 - Bronchitis, not specified as acute or chronic
J41 - Simple And Mucopurulent Chronic Bronchitis
J42 - Unspecified Chronic Bronchitis
J200 - Acute bronchitis due to Mycoplasma pneumoniae
J201 - Acute bronchitis due to Hemophilus influenzae
J202 - Acute bronchitis due to streptococcus
J203 - Acute bronchitis due to coxsackievirus
J204 - Acute bronchitis due to parainfluenza virus
J205 - Acute bronchitis due to respiratory syncytial virus

J206 - Acute bronchitis due to rhinovirus
J207 - Acute bronchitis due to echovirus
J208 - Acute bronchitis due to other specified organisms
J209 - Acute bronchitis, unspecified
J410 - Simple Chronic Bronchitis
J411 - Mucopurulent Chronic Bronchitis
J418 - Mixed Simple And Mucopurulent Chronic Bronchitis
J680 - Bronchitis and pneumonitis due to chemicals, gases, fumes and vapors

AIRWAY - Cough

7862 - Cough
49382 - Cough Variant Asthma
G4483 - Primary Cough Headache
J45991 - Cough Variant Asthma
R05 - Cough
R052 - Subacute cough
R053 - Chronic cough

AIRWAY - Serious Pulmonology Events

9601 - Acute Respiratory Failure With Hypoxia
51851 - Acute Respiratory Failure Following Trauma And Surgery
51853 - Acute And Chronic Respiratory Failure Following Trauma And Surgery
51881 - Acute Respiratory Failure
51883 - Chronic Respiratory Failure
51884 - Acute And Chronic Respiratory Failure
J80 - Acute Respiratory Distress Syndrome
J90 - Pleural Effusion, Not Elsewhere Classified
J96 - Respiratory Failure, Not Elsewhere Classified
J810 - Acute Pulmonary Edema
J960 - Acute Respiratory Failure (5.7k)
J961 - Chronic Respiratory Failure (3.1k)
J962 - Acute And Chronic Respiratory Failure (1.7k)
J969 - Respiratory Failure, Unspecified (1.6k)
J9582 - Postprocedural Respiratory Failure (2.1k)
J9600 - Acute Respiratory Failure, Unspecified Whether With Hypoxia Or Hypercapnia
J9601 - Acute Respiratory Failure With Hypoxia
J9602 - Acute Respiratory Failure With Hypercapnia
J9610 - Chronic Respiratory Failure, Unspecified Whether With Hypoxia Or Hypercapnia
J9611 - Chronic Respiratory Failure With Hypoxia
J9612 - Chronic Respiratory Failure With Hypercapnia
J9620 - Acute And Chronic Respiratory Failure, Unspecified Whether With Hypoxia Or Hypercapnia
J9621 - Acute And Chronic Respiratory Failure With Hypoxia
J9621 - Acute And Chronic Respiratory Failure With Hypoxia
J9622 - Acute And Chronic Respiratory Failure With Hypercapnia
J9690 - Respiratory Failure, Unspecified, Unspecified Whether With Hypoxia Or Hypercapnia
J9691 - Respiratory Failure, Unspecified With Hypoxia
J9692 - Respiratory Failure, Unspecified With Hypercapnia
J95821 - Acute Postprocedural Respiratory Failure
J95822 - Acute And Chronic Postprocedural Respiratory Failure

R0603 - Acute Respiratory Distress

AIRWAY - Pneumonia

481 - Pneumococcal Pneumonia [Streptococcus Pneumoniae Pneumonia]
486 - Pneumonia, Organism Unspecified
4829 - Bacterial Pneumonia, Unspecified
4838 - Pneumonia Due To Other Specified Organism
J13 - Pneumonia Due To Streptococcus Pneumoniae
J150 - Pneumonia Due To Klebsiella Pneumoniae
J151 - Pneumonia Due To Pseudomonas
J156 - Pneumonia Due To Other Gram-Negative Bacteria
J157 - Pneumonia Due To Mycoplasma Pneumoniae
J158 - Pneumonia Due To Other Specified Bacteria
J159 - Unspecified Bacterial Pneumonia
J168 - Pneumonia Due To Other Specified Infectious Organisms
J181 - Lobar Pneumonia, Unspecified Organism
J188 - Other Pneumonia, Unspecified Organism
J189 - Pneumonia, Unspecified Organism
J15212 - Pneumonia Due To Methicillin Resistant Staphylococcus Aureus
V1261 - Personal History Of Pneumonia (Recurrent)
Z8701 - Personal History Of Pneumonia (Recurrent)

AIRWAY - Upper Airway Infections

4658 - Acute Upper Respiratory Infections Of Other Multiple Sites
4659 - Acute Upper Respiratory Infections Of Unspecified Site
J069 - Acute Upper Respiratory Infection, Unspecified

CM - Congenital Malformations of Heart

I517 - Cardiomegaly
Q20 - Congenital Malformations Of Cardiac Chambers And Connections (3.5k)
Q21 - Congenital Malformations Of Cardiac Septa (3.4k)
Q24 - Other Congenital Malformations Of Heart (4.7k)
Q204 - Double Inlet Ventricle
Q208 - Other Congenital Malformations Of Cardiac Chambers And Connections
Q209 - Congenital Malformation Of Cardiac Chambers And Connections, Unspecified
Q210 - Ventricular Septal Defect
Q211 - Atrial Septal Defect
Q212 - Atrioventricular Septal Defect
Q213 - Tetralogy Of Fallot
Q214 - Aortopulmonary Septal Defect
Q218 - Other Congenital Malformations Of Cardiac Septa
Q219 - Congenital Malformation Of Cardiac Septum, Unspecified
Q238 - Other Congenital Malformations Of Aortic And Mitral Valves
Q240 - Dextrocardia
Q241 - Levocardia
Q242 - Cor Triatriatum (5.6k)
Q243 - Pulmonary Infundibular Stenosis
Q244 - Congenital Subaortic Stenosis
Q245 - Malformation Of Coronary Vessels
Q246 - Congenital Heart Block

Q248 - Other Specified Congenital Malformations Of Heart
Q249 - Congenital Malformation Of Heart, Unspecified

CM - Other Congenital Malformations

Q89 - Other Congenital Malformations, Not Elsewhere Classified (3.5k)
Q453 - Other Congenital Malformations Of Pancreas And Pancreatic Duct
Q638 - Other Specified Congenital Malformations Of Kidney
Q890 - Congenital Absence And Malformations Of Spleen (14)
Q891 - Congenital Malformations Of Adrenal Gland
Q892 - Congenital Malformations Of Other Endocrine Glands
Q897 - Multiple Congenital Malformations, Not Elsewhere Classified
Q898 - Other Specified Congenital Malformations
Q899 - Congenital Malformation, Unspecified
Q8901 - Asplenia (Congenital)
Q8909 - Congenital Malformations Of Spleen

CM - Situs Inversus

Q893 - Situs Inversus

DIAG. - Asthma

49300 - Extrinsic Asthma, Unspecified
49301 - Extrinsic Asthma With Status Asthmaticus
49302 - Extrinsic Asthma With  (Acute) Exacerbation
49310 - Intrinsic Asthma, Unspecified
49320 - Chronic Obstructive Asthma, Unspecified
49382 - Cough Variant Asthma
49390 - Asthma, Unspecified Type, Unspecified
49391 - Asthma, Unspecified Type, With Status Asthmaticus
49392 - Asthma, Unspecified Type, With  (Acute) Exacerbation
J455 - Severe Persistent Asthma (790)
J4520 - Mild Intermittent Asthma, Uncomplicated
J4521 - Mild Intermittent Asthma With  (Acute) Exacerbation
J4522 - Mild Intermittent Asthma With Status Asthmaticus
J4530 - Mild Persistent Asthma, Uncomplicated
J4531 - Mild Persistent Asthma With  (Acute) Exacerbation
J4540 - Moderate Persistent Asthma, Uncomplicated
J4541 - Moderate Persistent Asthma With  (Acute) Exacerbation
J4542 - Moderate Persistent Asthma With Status Asthmaticus
J4550 - Severe Persistent Asthma, Uncomplicated
J4550 - Severe Persistent Asthma, Uncomplicated
J4551 - Severe Persistent Asthma With  (Acute) Exacerbation
J4551 - Severe Persistent Asthma With (Acute) Exacerbation
J4552 - Severe Persistent Asthma With Status Asthmaticus
J45901 - Unspecified Asthma With  (Acute) Exacerbation
J45902 - Unspecified Asthma With Status Asthmaticus
J45909 - Unspecified Asthma, Uncomplicated
J45998 - Other Asthma
Z825 - Family History Of Asthma And Other Chronic Lower Respiratory Diseases

DIAG. - COVID-19

U071 - Covid-19

DIAG. - Cystic Fibrosis

27702 - Cystic Fibrosis With Pulmonary Manifestations
E84 - Cystic Fibrosis (1.5k)
E840 - Cystic Fibrosis With Pulmonary Manifestations
E849 - Cystic Fibrosis, Unspecified

DIAG. - Pulmonary Diagnoses

J984 - Other Disorders Of Lung
J988 - Other Specified Respiratory Disorders
J9809 - Other Diseases Of Bronchus, Not Elsewhere Classified
R918 - Other Nonspecific Abnormal Finding Of Lung Field
Z8709 - Personal History Of Other Diseases Of The Respiratory System

DIAG. - Q348 - Congenital Malf. Resp. Sys. (Specific)

Q348 - Other Specified Congenital Malformations Of Respiratory System (8.6k)

EAR - Otitis media

3813 - Other And Unspecified Chronic Nonsuppurative Otitis Media
3814 - Nonsuppurative Otitis Media, Not Specified As Acute Or Chronic
3821 - Chronic Tubotympanic Suppurative Otitis Media
3822 - Chronic Atticoantral Suppurative Otitis Media
3823 - Unspecified Chronic Suppurative Otitis Media
3829 - Unspecified Otitis Media
38100 - Acute Nonsuppurative Otitis Media, Unspecified
38101 - Acute Serous Otitis Media
38110 - Chronic Serous Otitis Media, Simple Or Unspecified
38119 - Other Chronic Serous Otitis Media
38120 - Chronic Mucoid Otitis Media, Simple Or Unspecified
38129 - Other Chronic Mucoid Otitis Media
38200 - Acute Suppurative Otitis Media Without Spontaneous Rupture Of Eardrum
H663X1 - Other Chronic Suppurative Otitis Media, Right Ear
H663X2 - Other Chronic Suppurative Otitis Media, Left Ear
H663X3 - Other Chronic Suppurative Otitis Media, Bilateral
H663X9 - Other Chronic Suppurative Otitis Media, Unspecified Ear
H6500 - Acute Serous Otitis Media, Unspecified Ear
H6501 - Acute Serous Otitis Media, Right Ear
H6502 - Acute Serous Otitis Media, Left Ear
H6503 - Acute Serous Otitis Media, Bilateral
H6520 - Chronic Serous Otitis Media, Unspecified Ear
H6521 - Chronic Serous Otitis Media, Right Ear
H6522 - Chronic Serous Otitis Media, Left Ear
H6523 - Chronic Serous Otitis Media, Bilateral
H6530 - Chronic Mucoid Otitis Media, Unspecified Ear
H6531 - Chronic Mucoid Otitis Media, Right Ear
H6532 - Chronic Mucoid Otitis Media, Left Ear
H6533 - Chronic Mucoid Otitis Media, Bilateral
H6590 - Unspecified Nonsuppurative Otitis Media, Unspecified Ear

H6591 - Unspecified Nonsuppurative Otitis Media, Right Ear
H6592 - Unspecified Nonsuppurative Otitis Media, Left Ear
H6593 - Unspecified Nonsuppurative Otitis Media, Bilateral
H6611 - Chronic Tubotympanic Suppurative Otitis Media, Right Ear
H6612 - Chronic Tubotympanic Suppurative Otitis Media, Left Ear
H6623 - Chronic Atticoantral Suppurative Otitis Media, Bilateral
H6641 - Suppurative Otitis Media, Unspecified, Right Ear
H6642 - Suppurative Otitis Media, Unspecified, Left Ear
H6643 - Suppurative Otitis Media, Unspecified, Bilateral
H6690 - Otitis Media, Unspecified, Unspecified Ear
H6691 - Otitis Media, Unspecified, Right Ear
H6692 - Otitis Media, Unspecified, Left Ear
H6693 - Otitis Media, Unspecified, Bilateral
H65191 - Other Acute Nonsuppurative Otitis Media, Right Ear
H65192 - Other Acute Nonsuppurative Otitis Media, Left Ear
H65193 - Other Acute Nonsuppurative Otitis Media, Bilateral
H65413 - Chronic Allergic Otitis Media, Bilateral
H65491 - Other Chronic Nonsuppurative Otitis Media, Right Ear
H65492 - Other Chronic Nonsuppurative Otitis Media, Left Ear
H65493 - Other Chronic Nonsuppurative Otitis Media, Bilateral
H65499 - Other Chronic Nonsuppurative Otitis Media, Unspecified Ear
H66001 - Acute Suppurative Otitis Media Without Spontaneous Rupture Of Ear Drum, Right Ear
H66002 - Acute Suppurative Otitis Media Without Spontaneous Rupture Of Ear Drum, Left Ear
H66003 - Acute Suppurative Otitis Media Without Spontaneous Rupture Of Ear Drum, Bilateral
H66006 - Acute Suppurative Otitis Media Without Spontaneous Rupture Of Ear Drum, Recurrent, Bilateral

EAR - Hearing Loss

389 - Hearing loss
3882 - Sudden hearing loss, unspecified
3890 - Conductive hearing loss
3891 - Sensorineural hearing loss
3892 - Mixed conductive and sensorineural hearing loss
3898 - Other specified forms of hearing loss
3899 - Unspecified hearing loss
31534 - Speech and language developmental delay due to hearing loss
38812 - Noise-induced hearing loss
38900 - Conductive hearing loss, unspecified
38901 - Conductive hearing loss, external ear
38902 - Conductive hearing loss, tympanic membrane
38903 - Conductive hearing loss, middle ear
38904 - Conductive hearing loss, inner ear
38905 - Conductive hearing loss, unilateral
38906 - Conductive hearing loss, bilateral
38908 - Conductive hearing loss of combined types
38910 - Sensorineural hearing loss, unspecified
38911 - Sensory hearing loss, bilateral
38912 - Neural hearing loss, bilateral
38913 - Neural hearing loss, unilateral
38914 - Central hearing loss
38915 - Sensorineural hearing loss, unilateral
38916 - Sensorineural hearing loss, asymmetrical

38917 - Sensory hearing loss, unilateral
38918 - Sensorineural hearing loss, bilateral
38920 - Mixed hearing loss, unspecified
38921 - Mixed hearing loss, unilateral
38922 - Mixed hearing loss, bilateral
F804 - Speech and language development delay due to hearing loss
H90 - Conductive and sensorineural hearing loss
H90A - Conductive and sensorineural hearing loss with restricted hearing on the contralateral side
H90A1 - Conductive hearing loss, unilateral, with restricted hearing on the contralateral side
H90A2 - Sensorineural hearing loss, unilateral, with restricted hearing on the contralateral side
H90A3 - Mixed conductive and sensorineural hearing loss, unilateral with restricted hearing on the contralateral side
H90A11 - Conductive hearing loss, unilateral, right ear with restricted hearing on the contralateral side
H90A12 - Conductive hearing loss, unilateral, left ear with restricted hearing on the contralateral side
H90A21 - Sensorineural hearing loss, unilateral, right ear, with restricted hearing on the contralateral side
H90A22 - Sensorineural hearing loss, unilateral, left ear, with restricted hearing on the contralateral side
H90A31 - Mixed conductive and sensorineural hearing loss, unilateral, right ear with restricted hearing on the contralateral side
H90A32 - Mixed conductive and sensorineural hearing loss, unilateral, left ear with restricted hearing on the contralateral side
H91 - Other and unspecified hearing loss
H900 - Conductive hearing loss, bilateral
H901 - Conductive hearing loss, unilateral with unrestricted hearing on the contralateral side
H902 - Conductive hearing loss, unspecified
H903 - Sensorineural hearing loss, bilateral
H904 - Sensorineural hearing loss, unilateral with unrestricted hearing on the contralateral side
H905 - Unspecified sensorineural hearing loss
H906 - Mixed conductive and sensorineural hearing loss, bilateral
H907 - Mixed conductive and sensorineural hearing loss, unilateral with unrestricted hearing on the contralateral side
H908 - Mixed conductive and sensorineural hearing loss, unspecified
H910 - Ototoxic hearing loss
H912 - Sudden idiopathic hearing loss
H918 - Other specified hearing loss
H918X - Other specified hearing loss
H918X1 - Other specified hearing loss, right ear
H918X2 - Other specified hearing loss, left ear
H918X3 - Other specified hearing loss, bilateral
H918X9 - Other specified hearing loss, unspecified ear
H919 - Unspecified hearing loss
H9011 - Conductive hearing loss, unilateral, right ear, with unrestricted hearing on the contralateral side
H9012 - Conductive hearing loss, unilateral, left ear, with unrestricted hearing on the contralateral side
H9041 - Sensorineural hearing loss, unilateral, right ear, with unrestricted hearing on the contralateral side
H9042 - Sensorineural hearing loss, unilateral, left ear, with unrestricted hearing on the contralateral side
H9071 - Mixed conductive and sensorineural hearing loss, unilateral, right ear, with unrestricted hearing on the contralateral side
H9072 - Mixed conductive and sensorineural hearing loss, unilateral, left ear, with unrestricted hearing on the contralateral side
H9101 - Ototoxic hearing loss, right ear
H9102 - Ototoxic hearing loss, left ear
H9103 - Ototoxic hearing loss, bilateral
H9109 - Ototoxic hearing loss, unspecified ear

H9120 - Sudden idiopathic hearing loss, unspecified ear
H9121 - Sudden idiopathic hearing loss, right ear
H9122 - Sudden idiopathic hearing loss, left ear
H9123 - Sudden idiopathic hearing loss, bilateral
H9190 - Unspecified hearing loss, unspecified ear
H9191 - Unspecified hearing loss, right ear
H9192 - Unspecified hearing loss, left ear
H9193 - Unspecified hearing loss, bilateral
P096 - Abnormal findings on neonatal screening for neonatal hearing loss
V192 - Family history of deafness or hearing loss
Z822 - Family history of deafness and hearing loss

EAR - Otorrhea

38860 - Otorrhea, Unspecified
38861 - Cerebrospinal Fluid Otorrhea (5.9k)
38869 - Other Otorrhea
H921 - Otorrhea (460)
H9210 - Otorrhea, Unspecified Ear
H9211 - Otorrhea, Right Ear
H9212 - Otorrhea, Left Ear
H9213 - Otorrhea, Bilateral

FERTILITY - Infertility

6069 - Male Infertility, Unspecified
6282 - Infertility, Female, Of Tubal Origin
6288 - Infertility, Female, Of Other Specified Origin
6289 - Infertility, Female, Of Unspecified Origin
N46 - Male Infertility (3.8k)
N97 - Female Infertility (4.2k)
N468 - Other Male Infertility
N469 - Male Infertility, Unspecified
N971 - Female Infertility Of Tubal Origin
N978 - Female Infertility Of Other Origin
N979 - Female Infertility, Unspecified

IMMUNE SYSTEM - Immunodeficiency

042 - Human Immunodeficiency Virus [Hiv] Disease
07953 - Human Immunodeficiency Virus, Type 2 [Hiv-2]
27901 - Selective Iga Immunodeficiency
27906 - Common Variable Immunodeficiency
27910 - Immunodeficiency With Predominant T-Cell Defect, Unspecified
79571 - Nonspecific Serologic Evidence Of Human Immunodeficiency Virus [Hiv]
B20 - Human Immunodeficiency Virus [Hiv] Disease
B9735 - Human Immunodeficiency Virus, Type 2 [Hiv 2] As The Cause Of Diseases Classified Elsewhere
D809 - Immunodeficiency With Predominantly Antibody Defects, Unspecified
D819 - Combined Immunodeficiency, Unspecified
D823 - Immunodeficiency Following Hereditary Defective Response To Epstein-Barr Virus
D830 - Common Variable Immunodeficiency With Predominant Abnormalities Of B-Cell Numbers And Function
D831 - Common Variable Immunodeficiency With Predominant Immunoregulatory T-Cell Disorders

D839 - Common Variable Immunodeficiency, Unspecified
D849 - Immunodeficiency, Unspecified
D8481 - Immunodeficiency Due To Conditions Classified Elsewhere
D84821 - Immunodeficiency Due To Drugs
D84822 - Immunodeficiency Due To External Causes
O9872 - Human Immunodeficiency Virus [Hiv] Disease Complicating Childbirth
O98712 - Human Immunodeficiency Virus [Hiv] Disease Complicating Pregnancy, Second Trimester
O98713 - Human Immunodeficiency Virus [Hiv] Disease Complicating Pregnancy, Third Trimester
O98719 - Human Immunodeficiency Virus [Hiv] Disease Complicating Pregnancy, Unspecified Trimester (9.6k)
R75 - Inconclusive Laboratory Evidence Of Human Immunodeficiency Virus [Hiv]
V08 - Asymptomatic Human Immunodeficiency Virus [Hiv] Infection Status
V6544 - Human Immunodeficiency Virus (Hiv) Counseling
Z21 - Asymptomatic Human Immunodeficiency Virus [Hiv] Infection Status
Z114 - Encounter For Screening For Human Immunodeficiency Virus [Hiv]
Z206 - Contact With And (Suspected) Exposure To Human Immunodeficiency Virus [Hiv]
Z717 - Human Immunodeficiency Virus [Hiv] Counseling
Z830 - Family History Of Human Immunodeficiency Virus [Hiv] Disease

NOSE - Nasal Congestion

R0981 - Nasal Congestion

NOSE - Nasal Polyps

4719 - Unspecified Nasal Polyp
4710 - Polyp Of Nasal Cavity
J33 - Nasal Polyp (2.7k)
J330 - Polyp Of Nasal Cavity
J339 - Nasal Polyp, Unspecified

NOSE - Rhinitis

4720 - Chronic Rhinitis
J300 - Vasomotor Rhinitis
J31 - Chronic Rhinitis, Nasopharyngitis And Pharyngitis (3.9k)
J310 - Chronic Rhinitis

QOL - Depression or Anxiety

311 - Depressive Disorder, Not Elsewhere Classified
29620 - Major Depressive Affective Disorder, Single Episode, Unspecified
29630 - Major Depressive Affective Disorder, Recurrent Episode, Unspecified
29632 - Major Depressive Affective Disorder, Recurrent Episode, Moderate
29633 - Major Depressive Affective Disorder, Recurrent Episode, Severe, Without Mention Of Psychotic
30000 - Anxiety State, Unspecified
30002 - Generalized Anxiety Disorder
30009 - Other Anxiety States
30924 - Adjustment Disorder With Anxiety
30928 - Adjustment Disorder With Mixed Anxiety And Depressed Mood
30928 - Adjustment Disorder With Mixed Anxiety And Depressed Mood
F32A - Depression, unspecified
F064 - Anxiety Disorder Due To Known Physiological Condition

F320 - Major Depressive Disorder, Single Episode, Mild
F320 - Major Depressive Disorder, Single Episode, Mild
F321 - Major Depressive Disorder, Single Episode, Moderate
F322 - Major Depressive Disorder, Single Episode, Severe Without Psychotic Features
F325 - Major Depressive Disorder, Single Episode, In Full Remission
F329 - Major Depressive Disorder, Single Episode, Unspecified
F330 - Major Depressive Disorder, Recurrent, Mild
F331 - Major Depressive Disorder, Recurrent, Moderate
F332 - Major Depressive Disorder, Recurrent Severe Without Psychotic Features
F339 - Major Depressive Disorder, Recurrent, Unspecified
F410 - Panic Disorder [Episodic Paroxysmal Anxiety]
F411 - Generalized Anxiety Disorder
F413 - Other Mixed Anxiety Disorders
F418 - Other Specified Anxiety Disorders
F419 - Anxiety Disorder, Unspecified
F3289 - Other Specified Depressive Episodes
F3341 - Major Depressive Disorder, Recurrent, In Partial Remission
F4321 - Adjustment Disorder With Depressed Mood
F4321 - Adjustment Disorder With Depressed Mood
F4322 - Adjustment Disorder With Anxiety
F4323 - Adjustment Disorder With Mixed Anxiety And Depressed Mood
F4323 - Adjustment Disorder With Mixed Anxiety And Depressed Mood
V790 - [Icd10] Driver Of Bus Injured In Collision ... (Or) [Icd9] Screening For Depression...

SINUS - Chronic Sinusitis

4730 - Chronic Maxillary Sinusitis
4731 - Chronic Frontal Sinusitis
4732 - Chronic Ethmoidal Sinusitis
4733 - Chronic Sphenoidal Sinusitis
4738 - Other Chronic Sinusitis
4739 - Unspecified Sinusitis (Chronic)
J32 - Chronic Sinusitis (3.6k)
J320 - Chronic Maxillary Sinusitis
J321 - Chronic Frontal Sinusitis
J322 - Chronic Ethmoidal Sinusitis
J323 - Chronic Sphenoidal Sinusitis
J328 - Other Chronic Sinusitis
J329 - Chronic Sinusitis, Unspecified

#### PROCEDURE CATEGORIES

We did not denote procedure categories into groups. Procedure codes are assigned feature types in the same way

- PROC. - Acapella/Flutter: (COUNT)
- PROC. - Adenoidectomy: (COUNT)
- PROC. - BAHA: (COUNT)
- PROC. - Bronchoalveoloar lavage: (COVERAGE)
- PROC. - Chest Wall Manipulation: (COVERAGE)
- PROC. - Chest X-Rays: (COVERAGE)
- PROC. - Cochlear Implant Maintenance: (COVERAGE)
- PROC. - Cochlear Implant Placement: (COUNT)

- PROC. - Detection of Micro-organsms: (COVERAGE)
- PROC. - Diagnosic EM: (FIRST)
- PROC. - Endoscopic airway exam: (COVERAGE)
- PROC. - Genetic Testing: (COUNT)
- PROC. - Hearing aid Fitting + Maintenance: (COVERAGE)
- PROC. - Hospitalization: (COUNT)
- PROC. - IPV: (COUNT)
- PROC. - Lung Function Measurement: (COVERAGE)
- PROC. - Lung Transplant: (COUNT)
- PROC. - Mastoidectomy: (COUNT)
- PROC. - Myringotomy Tubes: (COUNT)
- PROC. - Nasal Biopsy: (COUNT)
- PROC. - Nitric Oxide Measurement: (COVERAGE)
- PROC. - Oscillatory positive expiratory pressure: (COUNT)
- PROC. - Sinus Surgery: (COVERAGE)
- PROC. - Sputum Induction: (COVERAGE)
- PROC. - Steroid Injection: (COVERAGE)
- PROC. - Sweat Chloride Test: (COUNT)
- PROC. - Tonsil surgery: (COUNT)
- PROC. - Tympanoplasty: (COUNT)

#### PROCEDURE CODES

PROC. - Acapella/Flutter

S8185 - Flutter device

PROC. - Adenoidectomy

283 - Tonsillectomy with adenoidectomy
286 - Adenoidectomy without tonsillectomy
287 - Control of hemorrhage after tonsillectomy and adenoidectomy
42830 - Removal of adenoids patient younger than age 12, initial procedure
42831 - Removal of adenoids patient age 12 or over, initial procedure
42835 - Removal of adenoids patient younger than age 12, secondary procedure
42836 - Removal of adenoids patient age 12 or over, secondary procedure

PROC. - BAHA

69710 - Implantation or replacement of temporal bone conduction hearing device
69711 - Removal or repair of temporal bone conduction hearing device

PROC. - Bronchoalveoloar lavage

0B9C00Z - Drainage of Right Upper Lung Lobe with Drainage Device, Open Approach
0B9C0ZX - Drainage of Right Upper Lung Lobe, Open Approach, Diagnostic
0B9C3ZX - Drainage of Right Upper Lung Lobe, Percutaneous Approach, Diagnostic
0B9C3ZZ - Drainage of Right Upper Lung Lobe, Percutaneous Approach
0B9C4ZX - Drainage of Right Upper Lung Lobe, Percutaneous Endoscopic Approach, Diagnostic
0B9C4ZZ - Drainage of Right Upper Lung Lobe, Percutaneous Endoscopic Approach
0B9C7ZX - Drainage of Right Upper Lung Lobe, Via Natural or Artificial Opening, Diagnostic
0B9C7ZZ - Drainage of Right Upper Lung Lobe, Via Natural or Artificial Opening
0B9C8ZX - Drainage of Right Upper Lung Lobe, Via Natural or Artificial Opening Endoscopic, Diagnostic

0B9C8ZZ - Drainage of Right Upper Lung Lobe, Via Natural or Artificial Opening Endoscopic
0B9C30Z - Drainage of Right Upper Lung Lobe with Drainage Device, Percutaneous Approach
0B9C40Z - Drainage of Right Upper Lung Lobe with Drainage Device, Percutaneous Endoscopic Approach
0B9C70Z - Drainage of Right Upper Lung Lobe with Drainage Device, Via Natural or Artificial Opening
0B9C80Z - Drainage of Right Upper Lung Lobe with Drainage Device, Via Natural or Artificial Opening Endoscopic
0B9D00Z - Drainage of Right Middle Lung Lobe with Drainage Device, Open Approach
0B9D0ZX - Drainage of Right Middle Lung Lobe, Open Approach, Diagnostic
0B9D0ZZ - Drainage of Right Middle Lung Lobe, Open Approach
0B9D3ZX - Drainage of Right Middle Lung Lobe, Percutaneous Approach, Diagnostic
0B9D3ZZ - Drainage of Right Middle Lung Lobe, Percutaneous Approach
0B9D4ZX - Drainage of Right Middle Lung Lobe, Percutaneous Endoscopic Approach, Diagnostic
0B9D7ZX - Drainage of Right Middle Lung Lobe, Via Natural or Artificial Opening, Diagnostic
0B9D7ZZ - Drainage of Right Middle Lung Lobe, Via Natural or Artificial Opening
0B9D8ZX - Drainage of Right Middle Lung Lobe, Via Natural or Artificial Opening Endoscopic, Diagnostic
0B9D8ZZ - Drainage of Right Middle Lung Lobe, Via Natural or Artificial Opening Endoscopic
0B9D30Z - Drainage of Right Middle Lung Lobe with Drainage Device, Percutaneous Approach
0B9D40Z - Drainage of Right Middle Lung Lobe with Drainage Device, Percutaneous Endoscopic Approach
0B9D70Z - Drainage of Right Middle Lung Lobe with Drainage Device, Via Natural or Artificial Opening
0B9D80Z - Drainage of Right Middle Lung Lobe with Drainage Device, Via Natural or Artificial Opening Endoscopic
0B9F00Z - Drainage of Right Lower Lung Lobe with Drainage Device, Open Approach
0B9F0ZX - Drainage of Right Lower Lung Lobe, Open Approach, Diagnostic
0B9F0ZZ - Drainage of Right Lower Lung Lobe, Open Approach
0B9F3ZX - Drainage of Right Lower Lung Lobe, Percutaneous Approach, Diagnostic
0B9F3ZZ - Drainage of Right Lower Lung Lobe, Percutaneous Approach
0B9F4ZX - Drainage of Right Lower Lung Lobe, Percutaneous Endoscopic Approach, Diagnostic
0B9F4ZZ - Drainage of Right Lower Lung Lobe, Percutaneous Endoscopic Approach
0B9F7ZX - Drainage of Right Lower Lung Lobe, Via Natural or Artificial Opening, Diagnostic
0B9F7ZZ - Drainage of Right Lower Lung Lobe, Via Natural or Artificial Opening
0B9F8ZX - Drainage of Right Lower Lung Lobe, Via Natural or Artificial Opening Endoscopic, Diagnostic
0B9F8ZZ - Drainage of Right Lower Lung Lobe, Via Natural or Artificial Opening Endoscopic
0B9F30Z - Drainage of Right Lower Lung Lobe with Drainage Device, Percutaneous Approach
0B9F40Z - Drainage of Right Lower Lung Lobe with Drainage Device, Percutaneous Endoscopic Approach
0B9F70Z - Drainage of Right Lower Lung Lobe with Drainage Device, Via Natural or Artificial Opening
0B9F80Z - Drainage of Right Lower Lung Lobe with Drainage Device, Via Natural or Artificial Opening Endoscopic
0B9G00Z - Drainage of Left Upper Lung Lobe with Drainage Device, Open Approach
0B9G0ZX - Drainage of Left Upper Lung Lobe, Open Approach, Diagnostic
0B9G0ZZ - Drainage of Left Upper Lung Lobe, Open Approach
0B9G3ZX - Drainage of Left Upper Lung Lobe, Percutaneous Approach, Diagnostic
0B9G3ZZ - Drainage of Left Upper Lung Lobe, Percutaneous Approach
0B9G4ZX - Drainage of Left Upper Lung Lobe, Percutaneous Endoscopic Approach, Diagnostic
0B9G4ZZ - Drainage of Left Upper Lung Lobe, Percutaneous Endoscopic Approach
0B9G7ZX - Drainage of Left Upper Lung Lobe, Via Natural or Artificial Opening, Diagnostic
0B9G7ZZ - Drainage of Left Upper Lung Lobe, Via Natural or Artificial Opening
0B9G8ZX - Drainage of Left Upper Lung Lobe, Via Natural or Artificial Opening Endoscopic, Diagnostic
0B9G8ZZ - Drainage of Left Upper Lung Lobe, Via Natural or Artificial Opening Endoscopic
0B9G30Z - Drainage of Left Upper Lung Lobe with Drainage Device, Percutaneous Approach
0B9G40Z - Drainage of Left Upper Lung Lobe with Drainage Device, Percutaneous Endoscopic Approach

0B9G70Z - Drainage of Left Upper Lung Lobe with Drainage Device, Via Natural or Artificial Opening
0B9G80Z - Drainage of Left Upper Lung Lobe with Drainage Device, Via Natural or Artificial Opening Endoscopic
0B9J00Z - Drainage of Left Lower Lung Lobe with Drainage Device, Open Approach
0B9J0ZX - Drainage of Left Lower Lung Lobe, Open Approach, Diagnostic
0B9J0ZZ - Drainage of Left Lower Lung Lobe, Open Approach
0B9J3ZX - Drainage of Left Lower Lung Lobe, Percutaneous Approach, Diagnostic
0B9J3ZZ - Drainage of Left Lower Lung Lobe, Percutaneous Approach
0B9J4ZX - Drainage of Left Lower Lung Lobe, Percutaneous Endoscopic Approach, Diagnostic
0B9J4ZZ - Drainage of Left Lower Lung Lobe, Percutaneous Endoscopic Approach
0B9J7ZX - Drainage of Left Lower Lung Lobe, Via Natural or Artificial Opening, Diagnostic
0B9J7ZZ - Drainage of Left Lower Lung Lobe, Via Natural or Artificial Opening
0B9J8ZX - Drainage of Left Lower Lung Lobe, Via Natural or Artificial Opening Endoscopic, Diagnostic
0B9J8ZZ - Drainage of Left Lower Lung Lobe, Via Natural or Artificial Opening Endoscopic
0B9J30Z - Drainage of Left Lower Lung Lobe with Drainage Device, Percutaneous Approach
0B9J40Z - Drainage of Left Lower Lung Lobe with Drainage Device, Percutaneous Endoscopic Approach
0B9J70Z - Drainage of Left Lower Lung Lobe with Drainage Device, Via Natural or Artificial Opening
0B9J80Z - Drainage of Left Lower Lung Lobe with Drainage Device, Via Natural or Artificial Opening Endoscopic
31624 - Irrigation and suction of lung airways to obtain cells using an endoscope
88106 - Cell examination of body fluid, simple filter method
88108 - Cell examination of specimen, concentration technique
88112 - Cell examination of specimen, selective cellular enhancement technique

PROC. - Cochlear Implant Placement

09HD05Z - Insertion of Single Channel Cochlear Prosthesis into Right Inner Ear, Open Approach
09HD06Z - Insertion of Multiple Channel Cochlear Prosthesis into Right Inner Ear, Open Approach
09HD35Z - Insertion of Single Channel Cochlear Prosthesis into Right Inner Ear, Percutaneous Approach
09HD36Z - Insertion of Multiple Channel Cochlear Prosthesis into Right Inner Ear, Percutaneous Approach
09HD45Z - Insertion of Single Channel Cochlear Prosthesis into Right Inner Ear, Percutaneous Endoscopic Approach
09HD46Z - Insertion of Multiple Channel Cochlear Prosthesis into Right Inner Ear, Percutaneous Endoscopic Approach
09HE05Z - Insertion of Single Channel Cochlear Prosthesis into Left Inner Ear, Open Approach
09HE06Z - Insertion of Multiple Channel Cochlear Prosthesis into Left Inner Ear, Open Approach
09HE35Z - Insertion of Single Channel Cochlear Prosthesis into Left Inner Ear, Percutaneous Approach
09HE36Z - Insertion of Multiple Channel Cochlear Prosthesis into Left Inner Ear, Percutaneous Approach
09HE45Z - Insertion of Single Channel Cochlear Prosthesis into Left Inner Ear, Percutaneous Endoscopic Approach
09HE46Z - Insertion of Multiple Channel Cochlear Prosthesis into Left Inner Ear, Percutaneous Endoscopic Approach
2096 - Implantation or replacement of cochlear prosthetic device, not otherwise specified
2097 - Implantation or replacement of cochlear prosthetic device, single channel
2098 - Implantation or replacement of cochlear prosthetic device, multiple channel
69714 - Temporal bone implantation of cochlear stimulating system, accessed through the skin
69715 - Removal of mastoid bone with implantation of cochlear stimulating system, accessed through the skin
69717 - Temporal bone replacement of cochlear stimulating system, accessed through the skin
69718 - Removal of mastoid bone with removal and replacement (accessed through the skin) of cochlear stimulating system
69930 - Implantation of cochlear device

92584 - Testing of nerve from ear to brain (cochlear)
92588 - Placement of ear probe for computerized cochlear assessment of repeated sounds with interpretation and report

PROC. - Cochlear Implant Maintenance

92601 - Analysis and programming of inner ear (cochlear) implant, patient younger than 7 years of age
92602 - Analysis and reprogramming of inner ear (cochlear) implant, patient younger than 7 years of age
92603 - Analysis and programming of inner ear (cochlear) implant, patient age 7 years or older
92604 - Analysis and reprogramming of inner ear (cochlear) implant, patient age 7 years or older
F0B - Physical Rehabilitation and Diagnostic Audiology, Rehabilitation, Cochlear Implant Treatment
F0BZ0KZ - Cochlear Implant Rehabilitation Treatment using Audiovisual Equipment
F0BZ0PZ - Cochlear Implant Rehabilitation Treatment using Computer
F0BZ0YZ - Cochlear Implant Rehabilitation Treatment using Other Equipment
F0BZ01Z - Cochlear Implant Rehabilitation Treatment using Audiometer
F0BZ02Z - Cochlear Implant Rehabilitation Treatment using Sound Field / Booth
F0BZ09Z - Cochlear Implant Rehabilitation Treatment using Cochlear Implant Equipment
F00Z19Z - Speech Threshold Assessment using Cochlear Implant Equipment
F00Z29Z - Speech/Word Recognition Assessment using Cochlear Implant Equipment
F00Z59Z - Synthetic Sentence Identification Assessment using Cochlear Implant Equipment
F13Z09Z - Hearing Screening Assessment using Cochlear Implant Equipment
F13ZP9Z - Aural Rehabilitation Status Assessment using Cochlear Implant Equipment
F14Z0KZ - Cochlear Implant Assessment using Audiovisual Equipment
F14Z0LZ - Cochlear Implant Assessment using Assistive Listening Equipment
F14Z0PZ - Cochlear Implant Assessment using Computer
F14Z0YZ - Cochlear Implant Assessment using Other Equipment
F14Z0ZZ - Cochlear Implant Assessment
F14Z01Z - Cochlear Implant Assessment using Audiometer
F14Z02Z - Cochlear Implant Assessment using Sound Field / Booth
F14Z03Z - Cochlear Implant Assessment using Tympanometer
F14Z04Z - Cochlear Implant Assessment using Electroacoustic Immitance / Acoustic Reflex Equipment
F14Z05Z - Cochlear Implant Assessment using Hearing Aid Selection / Fitting / Test Equipment
F14Z07Z - Cochlear Implant Assessment using Electrophysiologic Equipment
F14Z09Z - Cochlear Implant Assessment using Cochlear Implant Equipment
L8614 - Cochlear device, includes all internal and external components
L8615 - Headset/headpiece for use with cochlear implant device, replacement
L8616 - Microphone for use with cochlear implant device, replacement
L8617 - Transmitting coil for use with cochlear implant device, replacement
L8618 - Transmitter cable for use with cochlear implant device or auditory osseointegrated device, replacement
L8619 - Cochlear implant, external speech processor and controller, integrated system, replacement
L8621 - Zinc air battery for use with cochlear implant device and auditory osseointegrated sound processors, replacement, each
L8622 - Alkaline battery for use with cochlear implant device, any size, replacement, each
L8623 - Lithium ion battery for use with cochlear implant device speech processor, other than ear level, replacement, each
L8624 - Lithium ion battery for use with cochlear implant or auditory osseointegrated device speech processor, ear level, replacement, each
L8625 - External recharging system for battery for use with cochlear implant or auditory osseointegrated device, replacement only, each
L8627 - Cochlear implant, external speech processor, component, replacement
L8628 - Cochlear implant, external controller component, replacement
L8629 - Transmitting coil and cable, integrated, for use with cochlear implant device, replacement

V5273 - Assistive listening device, for use with cochlear implant

PROC. - Chest Wall Manipulation

94667 - Demonstration and/or evaluation of manual maneuvers to chest wall to assist movement of lung secretions
94668 - Manual maneuvers to chest wall to assist movement of lung secretions
94669 - Mechanical chest wall manipulation for improvement in lung function
A7025 - High Frequency Chest Wall Oscillation System Vest, Replacement For Use With Patient Owned Eq (430)
A7026 - High Frequency Chest Wall Oscillation System Hose, Replacement For Use With Patient Owned Eq (570)
E0483 - High frequency chest wall oscillation system, includes all accessories and supplies, each

PROC. - Chest X-Rays

71010 - X-Ray Of Chest, 1 View, Front
71020 - X-Ray Of Chest, 2 Views, Front And Side
71020 - X-Ray Of Chest, 2 Views, Front And Side
71021 - X-Ray Of Chest, 2 Views, Front And Side
71022 - X-Ray Of Chest, 2 Views, Front And Side
71030 - X-Ray Of Chest, Minimum Of 4 Views
71035 - X-Ray Of Chest, Special Views
71045 - X-Ray Of Chest, 1 View
71046 - X-Ray Of Chest, 2 Views
71047 - X-Ray Of Chest, 3 Views
71048 - X-Ray Of Chest, Minimum Of 4 Views
71101 - X-Ray Of Ribs On One Side Of Body Including The Chest, Minimum Of 3 Views
71111 - X-Ray Of Both Sides Of The Ribs Including The Chest, Minimum Of 4 Views
71250 - Diagnostic CT scan of chest
71260 - Diagnostic CT scan of chest with contrast
74022 - Complete X-Ray Study Of Abdomen With Single X-Ray Of Chest

PROC. - Diagnostic EM

88348 - Electron microscopy for diagnosis

PROC. - Endoscopic airway exam

3321 - Bronchoscopy through artificial stoma
3322 - Fiber-optic bronchoscopy
3323 - Other bronchoscopy
31622 - Diagnostic examination of lung airways using an endoscope
31623 - Examination of lung airways using an endoscope
31625 - Biopsy of lung airways using an endoscope
C9751 - Bronchoscopy, rigid or flexible, transbronchial ablation of lesion(s) by microwave energy, including fluoroscopic guidance, when performed, with computed tomography acquisition(s) and 3-d rendering, computer-assisted, image-guided navigation, and endobronchial ultrasound (ebus) guided transtracheal and/or transbronchial sampling (eg, aspiration[s]/biopsy[ies]) and all mediastinal and/or hilar lymph node stations or structures and therapeutic intervention(s)

PROC. - Genetic Testing

82016 - Chemical analysis for genetic disorder
82017 - Chemical test for genetic disorder
86352 - Analysis of cell function and analysis for genetic marker
88245 - Chromosome analysis for genetic defects, baseline Sister Chromatid Exchange (SCE), 20-25 cells
88261 - Chromosome analysis for genetic defects, count 5 cells
88262 - Chromosome analysis for genetic defects, count 15-20 cells
88264 - Chromosome analysis for genetic defects, analyze 20-25 cells
88271 - DNA testing for genetic defects
88273 - Chromosome analysis for genetic defects, analyze 10-30 cells
88274 - Chromosome analysis for genetic defects, analyze 25-99 cells
88280 - Chromosome analysis for genetic defects, additional karyotypes, each study
88285 - Chromosome analysis for genetic defects, additional cells counted, each study
88289 - Chromosome analysis for genetic defects, additional high resolution study
88291 - Interpretation and report of genetic testing
88365 - Genetic sequencing localization, initial procedure

PROC. - Hearing aid Fitting + Maintenance

2095 - Implantation of electromagnetic hearing device
9548 - Fitting of hearing aid
92590 - Hearing aid examination and selection of one ear
92591 - Hearing aid examination and selection of both ears
92592 - Check of hearing aid of one ear
92593 - Check of hearing aid of both ears
92594 - Assessment of hearing aid function for one ear
92595 - Assessment of hearing aid function for both ears
F0DZ1KZ - Monaural Hearing Aid Device Fitting using Audiovisual Equipment
F0DZ1LZ - Monaural Hearing Aid Device Fitting using Assistive Listening Equipment
F0DZ1ZZ - Monaural Hearing Aid Device Fitting
F0DZ2KZ - Binaural Hearing Aid Device Fitting using Audiovisual Equipment
F0DZ2LZ - Binaural Hearing Aid Device Fitting using Assistive Listening Equipment
F0DZ2ZZ - Binaural Hearing Aid Device Fitting
F0DZ05Z - Tinnitus Masker Device Fitting using Hearing Aid Selection / Fitting / Test Equipment
F0DZ11Z - Monaural Hearing Aid Device Fitting using Audiometer
F0DZ12Z - Monaural Hearing Aid Device Fitting using Sound Field / Booth
F0DZ15Z - Monaural Hearing Aid Device Fitting using Hearing Aid Selection / Fitting / Test Equipment
F0DZ21Z - Binaural Hearing Aid Device Fitting using Audiometer
F0DZ22Z - Binaural Hearing Aid Device Fitting using Sound Field / Booth
F0DZ25Z - Binaural Hearing Aid Device Fitting using Hearing Aid Selection / Fitting / Test Equipment
F0DZ55Z - Assistive Listening Device Device Fitting using Hearing Aid Selection / Fitting / Test Equipment
F14 - Physical Rehabilitation and Diagnostic Audiology, Diagnostic Audiology, Hearing Aid Assessment
F14Z2KZ - Monaural Hearing Aid Assessment using Audiovisual Equipment
F14Z2LZ - Monaural Hearing Aid Assessment using Assistive Listening Equipment
F14Z2PZ - Monaural Hearing Aid Assessment using Computer
F14Z2ZZ - Monaural Hearing Aid Assessment
F14Z3KZ - Binaural Hearing Aid Assessment using Audiovisual Equipment
F14Z3LZ - Binaural Hearing Aid Assessment using Assistive Listening Equipment
F14Z3PZ - Binaural Hearing Aid Assessment using Computer
F14Z3ZZ - Binaural Hearing Aid Assessment
F14Z05Z - Cochlear Implant Assessment using Hearing Aid Selection / Fitting / Test Equipment
F14Z6ZZ - Binaural Electroacoustic Hearing Aid Check Assessment

F14Z8ZZ - Monaural Electroacoustic Hearing Aid Check Assessment
F14Z15Z - Ear Canal Probe Microphone Assessment using Hearing Aid Selection / Fitting / Test Equipment
F14Z21Z - Monaural Hearing Aid Assessment using Audiometer
F14Z22Z - Monaural Hearing Aid Assessment using Sound Field / Booth
F14Z23Z - Monaural Hearing Aid Assessment using Tympanometer
F14Z24Z - Monaural Hearing Aid Assessment using Electroacoustic Immitance / Acoustic Reflex Equipment
F14Z25Z - Monaural Hearing Aid Assessment using Hearing Aid Selection / Fitting / Test Equipment
F14Z31Z - Binaural Hearing Aid Assessment using Audiometer
F14Z32Z - Binaural Hearing Aid Assessment using Sound Field / Booth
F14Z33Z - Binaural Hearing Aid Assessment using Tympanometer
F14Z34Z - Binaural Hearing Aid Assessment using Electroacoustic Immitance / Acoustic Reflex Equipment
F14Z35Z - Binaural Hearing Aid Assessment using Hearing Aid Selection / Fitting / Test Equipment
F14Z55Z - Sensory Aids Assessment using Hearing Aid Selection / Fitting / Test Equipment
F14Z65Z - Binaural Electroacoustic Hearing Aid Check Assessment using Hearing Aid Selection / Fitting / Test Equipment
F14Z85Z - Monaural Electroacoustic Hearing Aid Check Assessment using Hearing Aid Selection / Fitting / Test Equipment
F15Z75Z - Tinnitus Masker Assessment using Hearing Aid Selection / Fitting / Test Equipment
Level 3: V5030-V5060 - Monaural Hearing Aid
Level 3: V5120-V5267 - Hearing Services - Hearing Aids
S0618 - Audiometry for hearing aid evaluation to determine the level and degree of hearing loss
S2230 - Implantation of magnetic component of semi-implantable hearing device on ossicles in middle ear
V532 - Fitting and adjustment of hearing aid
V5010 - Assessment for hearing aid
V5011 - Fitting/orientation/checking of hearing aid
V5014 - Repair/modification of a hearing aid
V5030 - Hearing aid, monaural, body worn, air conduction
V5040 - Hearing aid, monaural, body worn, bone conduction
V5050 - Hearing aid, monaural, in the ear
V5060 - Hearing aid, monaural, behind the ear
V5090 - Dispensing fee, unspecified hearing aid
V5100 - Hearing aid, bilateral, body worn
V5170 - Hearing aid, cros, in the ear
V5171 - Hearing aid, contralateral routing device, monaural, in the ear (ite)
V5171 - Hearing aid, contralateral routing device, monaural, in the ear (ite)
V5172 - Hearing aid, contralateral routing device, monaural, in the canal (itc)
V5180 - Hearing aid, cros, behind the ear
V5181 - Hearing aid, contralateral routing device, monaural, behind the ear (bte)
V5190 - Hearing aid, contralateral routing, monaural, glasses
V5210 - Hearing aid, bicros, in the ear
V5211 - Hearing aid, contralateral routing system, binaural, ite/ite
V5212 - Hearing aid, contralateral routing system, binaural, ite/itc
V5213 - Hearing aid, contralateral routing system, binaural, ite/bte
V5214 - Hearing aid, contralateral routing system, binaural, itc/itc
V5215 - Hearing aid, contralateral routing system, binaural, itc/bte
V5220 - Hearing aid, bicros, behind the ear
V5221 - Hearing aid, contralateral routing system, binaural, bte/bte
V5230 - Hearing aid, contralateral routing system, binaural, glasses
V5241 - Dispensing fee, monaural hearing aid, any type

V5242 - Hearing aid, analog, monaural, cic (completely in the ear canal)
V5243 - Hearing aid, analog, monaural, itc (in the canal)
V5244 - Hearing aid, digitally programmable analog, monaural, cic
V5245 - Hearing aid, digitally programmable, analog, monaural, itc
V5246 - Hearing aid, digitally programmable analog, monaural, ite (in the ear)
V5247 - Hearing aid, digitally programmable analog, monaural, bte (behind the ear)
V5248 - Hearing aid, analog, binaural, cic
V5249 - Hearing aid, analog, binaural, itc
V5250 - Hearing aid, digitally programmable analog, binaural, cic
V5251 - Hearing aid, digitally programmable analog, binaural, itc
V5252 - Hearing aid, digitally programmable, binaural, ite
V5253 - Hearing aid, digitally programmable, binaural, bte
V5254 - Hearing aid, digital, monaural, cic
V5255 - Hearing aid, digital, monaural, itc
V5256 - Hearing aid, digital, monaural, ite
V5257 - Hearing aid, digital, monaural, bte
V5258 - Hearing aid, digital, binaural, cic
V5259 - Hearing aid, digital, binaural, itc
V5260 - Hearing aid, digital, binaural, ite
V5261 - Hearing aid, digital, binaural, bte
V5262 - Hearing aid, disposable, any type, monaural
V5263 - Hearing aid, disposable, any type, binaural
V5267 - Hearing aid or assistive listening device/supplies/accessories, not otherwise specified
V5298 - Hearing aid, not otherwise classified
V5336 - Repair/modification of augmentative communicative system or device (excludes adaptive hearing aid)
Z461 - Encounter for fitting and adjustment of hearing aid

PROC. - Hospitalization

99219 - Hospital observation care, typically 50 minutes
99223 - Initial hospital inpatient care, typically 70 minutes per day
99232 - Subsequent hospital inpatient care, typically 25 minutes per day
99283 - Emergency department visit, moderately severe problem
99284 - Emergency department visit, problem of high severity
99285 - Emergency department visit, problem with significant threat to life or function
99291 - Critical care delivery critically ill or injured patient, first 30-74 minutes
99472 - Subsequent inpatient hospital critical care of infant or young child, 29 days through 24 months of age, per day
G0378 - Hospital observation service, per hour

PROC. - IPV

E0481 - Intrapulmonary percussive ventilation system and related accessories

PROC. - Lung Function Measurement

82803 - Blood gases measurement
94010 - Measurement and graphic recording of total and timed exhaled air capacity
94060 - Measurement and graphic recording of the amount and speed of breathed air, before and following medication administration

94070 - Multiple measurements and graphic recordings of the amount and speed of breathed air, before and following medication administration
94150 - Measurement of largest amount of air exhaled from lungs
94375 - Respiratory diagnostic testing (flow volume loop)
94726 - Determination of lung volumes using plethysmography
94729 - Measurement of lung diffusing capacity
94760 - Measurement of oxygen saturation in blood using ear or finger device

PROC. - Lung Transplant

0BYC0Z0 - Transplantation of Right Upper Lung Lobe, Allogeneic, Open Approach
0BYC0Z1 - Transplantation of Right Upper Lung Lobe, Syngeneic, Open Approach
0BYC0Z2 - Transplantation of Right Upper Lung Lobe, Zooplastic, Open Approach
0BYD0Z0 - Transplantation of Right Middle Lung Lobe, Allogeneic, Open Approach
0BYD0Z1 - Transplantation of Right Middle Lung Lobe, Syngeneic, Open Approach
0BYD0Z2 - Transplantation of Right Middle Lung Lobe, Zooplastic, Open Approach
0BYF0Z0 - Transplantation of Right Lower Lung Lobe, Allogeneic, Open Approach
0BYF0Z1 - Transplantation of Right Lower Lung Lobe, Syngeneic, Open Approach
0BYF0Z2 - Transplantation of Right Lower Lung Lobe, Zooplastic, Open Approach
0BYG0Z0 - Transplantation of Left Upper Lung Lobe, Allogeneic, Open Approach
0BYG0Z1 - Transplantation of Left Upper Lung Lobe, Syngeneic, Open Approach
0BYG0Z2 - Transplantation of Left Upper Lung Lobe, Zooplastic, Open Approach
0BYH0Z0 - Transplantation of Lung Lingula, Allogeneic, Open Approach
0BYH0Z1 - Transplantation of Lung Lingula, Syngeneic, Open Approach
0BYH0Z2 - Transplantation of Lung Lingula, Zooplastic, Open Approach
0BYJ0Z0 - Transplantation of Left Lower Lung Lobe, Allogeneic, Open Approach
0BYJ0Z1 - Transplantation of Left Lower Lung Lobe, Syngeneic, Open Approach
0BYJ0Z2 - Transplantation of Left Lower Lung Lobe, Zooplastic, Open Approach
0BYK0Z0 - Transplantation of Right Lung, Allogeneic, Open Approach
0BYK0Z1 - Transplantation of Right Lung, Syngeneic, Open Approach
0BYK0Z2 - Transplantation of Right Lung, Zooplastic, Open Approach
0BYL0Z0 - Transplantation of Left Lung, Allogeneic, Open Approach
0BYL0Z1 - Transplantation of Left Lung, Syngeneic, Open Approach
0BYL0Z2 - Transplantation of Left Lung, Zooplastic, Open Approach
0BYM0Z0 - Transplantation of Bilateral Lungs, Allogeneic, Open Approach
0BYM0Z1 - Transplantation of Bilateral Lungs, Syngeneic, Open Approach
0BYM0Z2 - Transplantation of Bilateral Lungs, Zooplastic, Open Approach
32851 - Transplant of lung
32852 - Transplant of lung on heart-lung machine
32853 - Transplant of both lungs
32854 - Transplant of both lungs on heart-lung machine
33935 - Transplantation of donor heart and lung
S2152 - Solid organ(s), complete or segmental, single organ or combination of organs; deceased or living donor(s), procurement, transplantation, and related complications; including: drugs; supplies; hospitalization with outpatient follow-up; medical/surgical, diagnostic, emergency, and rehabilitative services, and the number of days of pre- and post-transplant care in the global definition

PROC. - Mastoidectomy

202 - Incision of mastoid and middle ear
204 - Mastoidectomy
2021 - Incision of mastoid
2041 - Simple mastoidectomy

2042 - Radical mastoidectomy
2049 - Other mastoidectomy
2092 - Revision of mastoidectomy
69220 - Removal of skin debris and drainage of mastoid cavity, simple
69222 - Removal of skin debris and drainage of mastoid cavity, complex
69501 - Incision of mastoid bone
69502 - Removal of mastoid bone
69505 - Removal of mastoid bone including removal of growth of middle ear
69511 - Removal of mastoid bone including removal of growth and bone of middle ear
69530 - Removal of portion of temporal bone including removal of mastoid bone
69552 - Removal of growth of external ear through mastoid bone
69601 - Revision of previous mastoid surgery with removal of remaining mastoid bone
69602 - Revision of previous mastoid surgery, modified radical procedure
69603 - Revision of previous mastoid surgery, radical procedure
69604 - Revision of previous mastoid surgery and ear drum
69605 - Revision of previous mastoid surgery
69635 - Repair of eardrum and ear canal with incision of mastoid bone
69636 - Repair of eardrum, ear canal, and bones with incision of mastoid bone
69637 - Repair of eardrum, ear canal, and bones with insertion of prosthesis with opening of mastoid
69641 - Repair of eardrum and ear canal with removal of mastoid bone, complex
69642 - Repair of eardrum, ear canal and bones with removal of mastoid bone, simple
69643 - Repair of eardrum and ear canal with removal of mastoid bone, simple
69644 - Repair of eardrum, ear canal and bones with removal of mastoid bone, with intact canal wall
69645 - Repair of eardrum and ear canal with removal of mastoid bone, extensive or radical
69646 - Repair of eardrum, ear canal and bones with removal of mastoid bone, extensive or radical
69715 - Removal of mastoid bone with implantation of cochlear stimulating system, accessed through the skin
69718 - Removal of mastoid bone with removal and replacement (accessed through the skin) of cochlear stimulating system
69910 - Removal of inner ear canal and removal of mastoid bone

PROC. - Myringotomy Tubes

2001 - Myringotomy With Insertion Of Tube
69436 - Incision of eardrum with insertion of eardrum tube under general anesthesia
Z4582 - Encounter For Adjustment Or Removal Of Myringotomy Device (Stent) (Tube)
Z9622 - Myringotomy Tube(S) Status

PROC. - Nasal Biopsy

30100 - Biopsy of lining of nose
31237 - Biopsy or removal of nasal polyp or tissue using an endoscope

PROC. - Nitric Oxide Measurement

95012 - Measurement of inhaled nitric oxide gas

PROC. - Oscillatory positive expiratory pressure

E0484 - Oscillatory positive expiratory pressure device, non-electric, any type, each

PROC. - Sinus Surgery

22 - Operations on nasal sinuses
0121 - Incision and drainage of cranial sinus
00160 - Anesthesia for procedure on nose and sinus
00162 - Anesthesia for surgery of nose and sinus
00164 - Anesthesia for soft tissue biopsy on nose and sinus
220 - Aspiration and lavage of nasal sinus
221 - Diagnostic procedures on nasal sinus
224 - Frontal sinusotomy and sinusectomy
225 - Other nasal sinusotomy
226 - Other nasal sinusectomy
227 - Repair of nasal sinus
229 - Other operations on nasal sinuses
0406T - Examination of nasal passage and sinus using an endoscope with placement of implant
0407T - Examination of nasal passage and sinus using an endoscope with placement of implant, biopsy and removal of polyps
1821 - Excision of preauricular sinus
2200 - Aspiration and lavage of nasal sinus, not otherwise specified
2201 - Puncture of nasal sinus for aspiration or lavage
2202 - Aspiration or lavage of nasal sinus through natural ostium
2211 - Closed [endoscopic] [needle] biopsy of nasal sinus
2212 - Open biopsy of nasal sinus
2219 - Other diagnostic procedures on nasal sinuses
2241 - Frontal sinusotomy
2242 - Frontal sinusectomy
2253 - Incision of multiple nasal sinuses
2261 - Excision of lesion of maxillary sinus with Caldwell-Luc approach
2262 - Excision of lesion of maxillary sinus with other approach
2271 - Closure of nasal sinus fistula
2279 - Other repair of nasal sinus
8603 - Incision of pilonidal sinus or cyst
8621 - Excision of pilonidal cyst or sinus
8935 - Transillumination of nasal sinuses
21139 - Repair of frontal sinus through forehead
21343 - Open treatment of frontal sinus fracture
21344 - Open treatment of depressed frontal sinus fracture
30210 - Irrigation and drainage of sinus
30580 - Repair of abnormal drainage tract between two nasal sinuses
31000 - Irrigation of nasal sinus (maxillary)
31002 - Irrigation of nasal sinus (sphenoid)
31020 - Incision of nasal (maxillary) sinus through the nose
31030 - Create a window into the nasal (maxillary) sinus
31032 - Removal of nasal sinus growths
31040 - Incision through sinus at cheek bone to reach nerves and blood vessels
31050 - Incision of nasal (sphenoid) sinus
31070 - Incision of nasal (frontal) sinus
31075 - Incision of nasal sinus of one side of face
31080 - Insertion of material to stop growth of nasal sinus lining without a bone flap done through an incision below the eyebrow
31081 - Insertion of material to stop growth of nasal sinus lining without a bone flap done through an incision through the forehead
31084 - Insertion of material to stop growth of nasal sinus lining with a bone flap done through an incision below the eyebrow

31085 - Insertion of material to stop growth of nasal sinus lining with a bone flap done through an incision through the forehead
31086 - Incision under the eyebrow to drain the nasal (frontal) sinus with placement of bone graft
31087 - Incision through the forehead to drain the nasal (frontal) sinus with placement of bone graft
31090 - Removal of diseased tissue or growths in multiple nasal sinuses on one side of face
31200 - Partial removal of nasal sinus
31201 - Removal of nasal sinus from within the nose passage
31205 - Removal of nasal sinus from outside the nose passage
31225 - Removal of nasal sinus
31230 - Removal of nasal sinus and eye bone
31233 - Examination of nasal passage and sinus above teeth (maxillary sinus) using endoscope
31235 - Examination of nasal passage and sinus above eyes (sphenoid sinus) using endoscope
31253 - Complete examination of nose and sinuses using an endoscope
31254 - Partial removal of nasal sinus using an endoscope
31255 - Complete removal of nasal sinus using an endoscope
31256 - Incision of nasal (maxillary) sinus using an endoscope
31257 - Complete examination of nose and sinuses and removal of nasal sinus using an endoscope
31259 - Removal of tissue from sphenoid sinus using an endoscope
31267 - Removal of nasal sinus tissue using an endoscope, maxillary sinus
31276 - Exploration of nasal sinus using an endoscope
31287 - Incision of nasal (sphenoid) sinus using an endoscope
31288 - Removal of nasal sinus tissue using an endoscope, sphenoid sinus
31290 - Repair of leak of brain and spinal fluid from sinus behind bridge of nose using endoscope
31291 - Repair of leak of brain and spinal fluid from sinus behind eyes using endoscope
31295 - Dilation of maxillary sinus in the nose using an endoscope
31296 - Dilation of frontal sinus in the nose using an endoscope
31297 - Dilation of sphenoid sinus in the nose using an endoscope
31298 - Dilation of sphenoid and frontal sinus in the nose using an endoscope
31299 - Accessory sinus procedure
42260 - Repair of abnormal connection from nasal sinus to skin surface
61580 - Removal of nasal sinuses to approach brain lesion without the removal of the maxilla or eyeball
61581 - Removal of nasal sinuses to approach brain lesion with the removal of the maxilla or eyeball
61598 - Removal of skull to approach lesion or defect at skull base with tying of sinus
C9771 - Nasal/sinus endoscopy, cryoablation nasal tissue(s) and/or nerve(s), unilateral or bilateral
G2097 - Episodes where the patient had a competing diagnosis on or within three days after the episode date (e.g., intestinal infection, pertussis, bacterial infection, lyme disease, otitis media, acute sinusitis, chronic sinusitis, infection of the adenoids, prostatitis, cellulitis, mastoiditis, or bone infections, acute lymphadenitis, impetigo, skin staph infections, pneumonia/gonococcal infections, venereal disease (syphilis, chlamydia, inflammatory diseases [female reproductive organs]), infections of the kidney, cystitis or uti)
G9350 - Ct scan of the paranasal sinuses not ordered at the time of diagnosis or received within 28 days after date of diagnosis
G9354 - One ct scan or no ct scan of the paranasal sinuses ordered within 90 days after the date of diagnosis
S2342 - Nasal endoscopy for post-operative debridement following functional endoscopic sinus surgery, nasal and/or sinus cavity(s), unilateral or bilateral
S9024 - Paranasal sinus ultrasound

PROC. - Detection of Micro-organsms

31624 - Irrigation and suction of lung airways to obtain cells using an endoscope
86140 - Measurement C-reactive protein for detection of infection or inflammation
86317 - Detection of infectious agent antibody, quantitative

87015 - Concentration of specimen for infectious agents
87040 - Bacterial blood culture
87070 - Bacterial culture, any other source except urine, blood or stool, aerobic ,
87071 - Bacterial culture and colony count
87075 - Bacterial culture, any source, except blood, anaerobic
87077 - Bacterial culture for aerobic isolates ,
87081 - Screening test for pathogenic organisms
87086 - Bacterial colony count, urine
87116 - Culture for acid-fast bacilli ,
87205 - Special Gram or Giemsa stain for microorganism ,
87206 - Special fluorescent and/or acid fast stain for microorganism
87299 - Detection test by immunofluorescent technique for organism
87486 - Detection test by nucleic acid for Chlamydia pneumoniae, amplified probe technique
87556 - Detection test by nucleic acid for Mycobacteria tuberculosis (TB bacteria), amplified probe technique
87581 - Detection test by nucleic acid for Mycoplasma pneumoniae (bacteria), amplified probe technique
87632 - Detection test by nucleic acid for multiple types of respiratory virus, multiple types or subtypes, 6-11 targets
87633 - Detection test by nucleic acid for multiple types of respiratory virus, multiple types or subtypes, 12-25 targets
87798 - Detection test by nucleic acid for organism, amplified probe technique
87804 - Detection test by immunoassay for influenza virus
87880 - Strep test by immunoassay for Streptococcus
88305 - Pathology examination of tissue using a microscope, intermediate complexity
88312 - Special stained specimen slides to identify organisms including interpretation and report
U0003 - Infectious agent detection by nucleic acid (dna or rna); severe acute respiratory syndrome coronavirus 2 (sars-cov-2) (coronavirus disease [covid-19]), amplified probe technique, making use of high throughput technologies as described by cms-2020-01-r
U0005 - Infectious agent detection by nucleic acid (dna or rna); severe acute respiratory syndrome coronavirus 2 (sars-cov-2) (coronavirus disease [covid-19]), amplified probe technique, cdc or non-cdc, making use of high throughput technologies, completed within 2 calendar days from date of specimen collection (list separately in addition to either hcpcs code u0003 or u0004) as described by cms-2020-01-r2

PROC. - Sputum Induction

89220 - Sputum Specimen Collection
94640 - Respiratory inhaled pressure or nonpressure treatment to relieve airway obstruction or for sputum specimen

PROC. - Steroid Injection

J1100 - Injection, dexamethasone sodium phosphate, 1 mg
J2704 - Injection, propofol, 10 mg

PROC. - Sweat Chloride Test

82438 - Chloride level
89230 - Sweat collection

PROC. - Tonsil surgery

0C5P0ZZ - Destruction of Tonsils, Open Approach

0C5P3ZZ - Destruction of Tonsils, Percutaneous Approach
0C5PXZZ - Destruction of Tonsils, External Approach
0C9P00Z - Drainage of Tonsils with Drainage Device, Open Approach
0C9P0ZX - Drainage of Tonsils, Open Approach, Diagnostic
0C9P0ZZ - Drainage of Tonsils, Open Approach
0C9P3ZX - Drainage of Tonsils, Percutaneous Approach, Diagnostic
0C9P3ZZ - Drainage of Tonsils, Percutaneous Approach
0C9P30Z - Drainage of Tonsils with Drainage Device, Percutaneous Approach
0C9PX0Z - Drainage of Tonsils with Drainage Device, External Approach
0C9PXZX - Drainage of Tonsils, External Approach, Diagnostic
0C9PXZZ - Drainage of Tonsils, External Approach
0CBP0ZX - Excision of Tonsils, Open Approach, Diagnostic
0CBP0ZZ - Excision of Tonsils, Open Approach
0CBP3ZX - Excision of Tonsils, Percutaneous Approach, Diagnostic
0CBP3ZZ - Excision of Tonsils, Percutaneous Approach
0CBPXZX - Excision of Tonsils, External Approach, Diagnostic
0CBPXZZ - Excision of Tonsils, External Approach
0CCP0ZZ - Extirpation of Matter from Tonsils, Open Approach
0CCP3ZZ - Extirpation of Matter from Tonsils, Percutaneous Approach
0CCPXZZ - Extirpation of Matter from Tonsils, External Approach
0CNP0ZZ - Release Tonsils, Open Approach
0CNP3ZZ - Release Tonsils, Percutaneous Approach
0CNPXZZ - Release Tonsils, External Approach
0CQP0ZZ - Repair Tonsils, Open Approach
0CQP3ZZ - Repair Tonsils, Percutaneous Approach
0CQPXZZ - Repair Tonsils, External Approach
0CTP0ZZ - Resection of Tonsils, Open Approach
0CTPXZZ - Resection of Tonsils, External Approach
42700 - Drainage of tonsil abscess
42820 - Removal of tonsils and adenoid glands patient younger than age 12
42821 - Removal of tonsils and adenoid glands patient age 12 or over
42825 - Removal of tonsils patient younger than age 12
42826 - Removal of tonsils patient age 12 or over
42842 - Removal of tonsils, tissue, muscle, and bone, without closure
42844 - Removal of tonsils, tissue, muscle, and bone, closure with local flap
42845 - Removal of tonsils, tissue, muscle, and bone, closure with other flap
42860 - Removal of remaining tonsil tissue

PROC. - Tympanoplasty

69633 - Repair of eardrum, ear canal, and bones with insertion of prosthesis, without mastoidectomy
69635 - Repair of eardrum and ear canal with incision of mastoid bone
69636 - Repair of eardrum, ear canal, and bones with incision of mastoid bone
69637 - Repair of eardrum, ear canal, and bones with insertion of prosthesis with opening of mastoid
69642 - Repair of eardrum, ear canal and bones with removal of mastoid bone, simple
69643 - Repair of eardrum and ear canal with removal of mastoid bone, simple
69644 - Repair of eardrum, ear canal and bones with removal of mastoid bone, with intact canal wall
69646 - Repair of eardrum, ear canal and bones with removal of mastoid bone, extensive or radical

**DRUG CODES**

We enumerated all codes for diagnoses and procedures but had to use regular expression (regex) searches to specify drugs since the number of separate codes in prescriptions was too large. Note that these are standard python regular expressions converted to lowercase.

These were grouped into the following categories:

- DRUG - Anti-inflammatory (Non-Steroid)
- DRUG - Anti-inflammatory (Steroid)
- DRUG - Antibiotics
- DRUG - Antidepressant
- DRUG - Antifungals
- DRUG - Antihistamine
- DRUG - Bronchodilators
- DRUG - Decongestant
- DRUG - Expectorant
- DRUG - Inhaled Hypertonic Saline
- DRUG - Mucolytics
- DRUG - Prophylactic Azithromycin

DRUG - Antibiotics

(inject|oral).*(claforan|duriflex|fortaz|keflex|rocephin|suprax|vantin)
(tobramycin|plazomicin|streptomycin|gentamicin|amikacin) .* injection
amikacin.*inhalation suspension
amikacin.*injectable solution
amoxicillin.*(oral|inject)
ampicillin.*(oral|inject)
augmentin.*(oral|inject)
azithromycin.*(oral|inject)
aztreonam .* (injection|inhalation solution)
carbenicillin
ceftazidime.*inject
ciprofloxacin .* (oral|injection)
ciprofloxacin .* otic suspension
clarithromycin.*(oral|inject)
clindamycin.*(oral|inject)
colistin.*solution
doxycycline.*oral
erythromycin.*(oral|inject)
gatifloxacin
gentamicin.*(irrigation|injectable)
imipenem
levofloxacin.*(oral|inject)
moxifloxacin.*(oral|inject)
mupirocin.*nasal
ofloxacin.*(ophthalmic),
otic suspension .*cortisporin
piperacillin.*inject
sulfamethoxazole.*trimethoprim.*inject.*
sulfamethoxazole.*trimethoprim.*oral
tetracycline.*oral
tobradex
tobramycin.*inhalation solution

tobramycin.*injectable solution
tobramycin.*podhaler
vancomycin.*inject

DRUG - Antidepressant

amitriptyline.*oral
bupropion.*oral
desvenlafaxine.*oral
duloxetine.*oral
fluoxetine.*oral
mirtazapine.*oral
nortriptyline.*oral
paroxetine.*oral

DRUG - Antifungals

amphotericin .* (oral|inject)
clotrimazole.*otic
fluconazole.*(oral|inject)
itraconazole.*(oral|inject)
ketoconazole .* (oral|otic)
voriconazole.*(oral|otic)

DRUG - Antihistamine

azelastine.*nasal
brompheniramine.*oral
cetirizine.*oral
chlorpheniramine.*oral
clemastine.*oral
diphenhydramine.*oral
exofenadine.*oral
levocetirizine.*oral
loratadine.*oral

DRUG - Anti-inflammatory (Steroid)

beclomethasone dipropionate .* inhaler
budesonide .* (inhalation|inhaler|nasal spray)
dexamethasone .* injection
fluticasone .* (inhalation|inhaler|nasal spray)
prednisolone .* oral
prednisone .* oral

DRUG - Anti-inflammatory (Non-Steroid)

ibuprofen .* oral
montelukast .* (oral|chewable)
naproxen .* oral

DRUG - Bronchodilators

aclidinium.*inhaler.
albuterol [48] mg extended release oral tablet
albuterol.*(inhaler|inhalation)
atrovent
formoterol
indacaterol
ipratropium.*(inhalation|inhaler)
levalbuterol
olodaerol
pirbuterol
salmeterol .* inhaler
salmeterol
terbutaline
theophylline.*oral (tablet|capsule)
tiotropium
umeclidinium
vilanterol

DRUG - Decongestant

(oxymetazoline|neosynephrine|xylometazoline|phenylephrin).*nasal
nasal.*afrin

DRUG - Expectorant

guaifenesin

DRUG - Mucolytics

acetylcysteine (5|25)00 mg effervescent oral tablet
acetylcysteine [12]00 mg/ml inhalation solution
acetylcysteine [56]00 mg oral (tablet|capsule)
bronchitol
dornase alfa 1 mg/ml inhalation solution
sodium chloride (4|5|6.5|6.9|7.4|9|9.5|10.5|11.5|20|21|26|26.5|30) mg/ml nasal (spray|solution)
sodium chloride 100 mg/ml inhalation solution
sodium chloride 30 mg/ml inhalation solution
sodium chloride 35 mg/ml inhalation solution
sodium chloride 60 mg/ml inhalation solution
sodium chloride 70 mg/ml inhalation solution
